# REVIVE-PEEP trial RESEARCH PROTOCOL

**DOI:** 10.64898/2026.02.26.26346617

**Authors:** Jeroen A. van Eijk, Judith ter Schure, Michiel Hulleman, Stephan A. Loer, Lothar A. Schwarte, Thijs Delnoij, Hans van Schuppen, Patrick Schober

## Abstract

**Rationale:** Cardiopulmonary resuscitation (CPR) aims to provide oxygen to vital organs through chest compressions and ventilations, until return of spontaneous circulation (ROSC) is achieved. A major barrier to effective oxygenation during CPR is atelectasis, which impairs gas exchange and results in hypoxemia—a condition strongly associated with decreased rates of ROSC and worsened neurological outcomes.

Positive end-expiratory pressure (PEEP) is routinely used in critical care to prevent atelectasis and improve oxygenation. During CPR, however, its use is inconsistent and subject of debate. This reflects long-standing theoretical concerns that PEEP may reduce venous return, lower cardiac output, and impair the chances of successful defibrillation. However, emerging experimental and observational data suggest that PEEP may actually improve oxygenation, cardiac output, oxygen delivery, and rates of ROSC during CPR.

We hypothesize that applying PEEP during CPR improves neurologically favorable survival. Given its simplicity, negligible cost, and widespread availability, PEEP has the potential to enhance outcomes from cardiac arrest in a scalable and resource-efficient manner.

**Objective:** To determine whether, during CPR with manual ventilation and after placement of a supraglottic airway device (SAD) or endotracheal tube (ETT), using a valve that generates 8 cm H_2_O PEEP compared with a sham valve providing zero end-expiratory pressure (ZEEP) improves neurological outcomes at discharge.

**Study design:** The REVIVE-PEEP trial is an investigator-initiated, pragmatic, registry-based, multicenter, parallel-group, triple-blind randomized controlled superiority clinical trial in the ARREST registry.

**Study population:** The target population consists of adult patients with an out-of-hospital cardiac arrest (OHCA) in whom advanced airway management is initiated, defined by placement of a SAD or ETT, followed by manual positive-pressure ventilation during CPR.

As randomization occurs at the start of CPR, initiation of advanced airway management is considered an intercurrent event and is used to define the modified intention-to-treat analysis population. Blinding ensures that analyses within this subset provide an unbiased estimate of the principal stratum estimand. The target sample size for the mITT subset is 2,400 patients.

**Intervention:** Participating ambulance services will use pre-assembled, 1:1 pre-randomized CPR kits containing a bag-valve-mask system with either a PEEP valve (8 cm H_2_O) or a sham valve (0 cm H_2_O; ZEEP), alongside standard advanced life support. The same resuscitator bag will be used throughout the resuscitation, both before and after advanced airway management.

**Main study parameters/endpoints:** The primary estimand is the difference in neurological outcome at hospital discharge, assessed by the utility-weighted modified Rankin Scale within the mITT subset. Key secondary outcomes include ROSC, 30-day survival and quality of life at 6 months.

**Nature and extent of the burden and risks associated with participation, benefit and group relatedness:** This study’s intervention involves only a minor adjustment to ventilation management during cardiac arrest and requires no additional procedures. The study design imposes no additional clinical tasks on ambulance professionals during resuscitation, allowing them to maintain full focus on patient care; the only study-related action is recording the study device number in the case report form after the resuscitation attempt.

The risks associated with participation are minimal. There may be a negligible increase in thoracic impedance that theoretically could influence defibrillation; however, modern defibrillators automatically adjust delivered energy based on pre-defibrillation thoracic impedance. Leakage around the SAD is a known issue that could reduce ventilation efficiency and may be exacerbated by the intervention, but existing guidelines allow for switching to a 30:2 compression:ventilation ratio in such cases. Lastly, although higher levels of PEEP have been associated with reduced cardiac output during CPR, the PEEP level used in this study remains under the maximum tolerated dose.

Potential benefits include improved oxygenation and ventilation, reduced afterload, and improved cardiopulmonary interactions, which may increase the likelihood of successful resuscitation and favorable neurological outcomes. Improved oxygenation may also increase the likelihood of successful defibrillation.

## 1. INTRODUCTION AND RATIONALE

### 1.1 Burden and unmet need

OHCA remains a significant health burden globally, with approximately four million cases annually, including 17,000 in the Netherlands.^1^ Although 30-day survival in the Netherlands has doubled—from 9% in the 1990s to about 20% today—thanks to initiatives like community first responder programs, AED deployment, and rapid ambulance responses, overall survival remains far from satisfactory.^2^ Moreover, survival has stagnated over the past decade (Figure 1), highlighting the urgent need for novel, evidence-based interventions to further improve patient outcomes.

**Figure 1.**
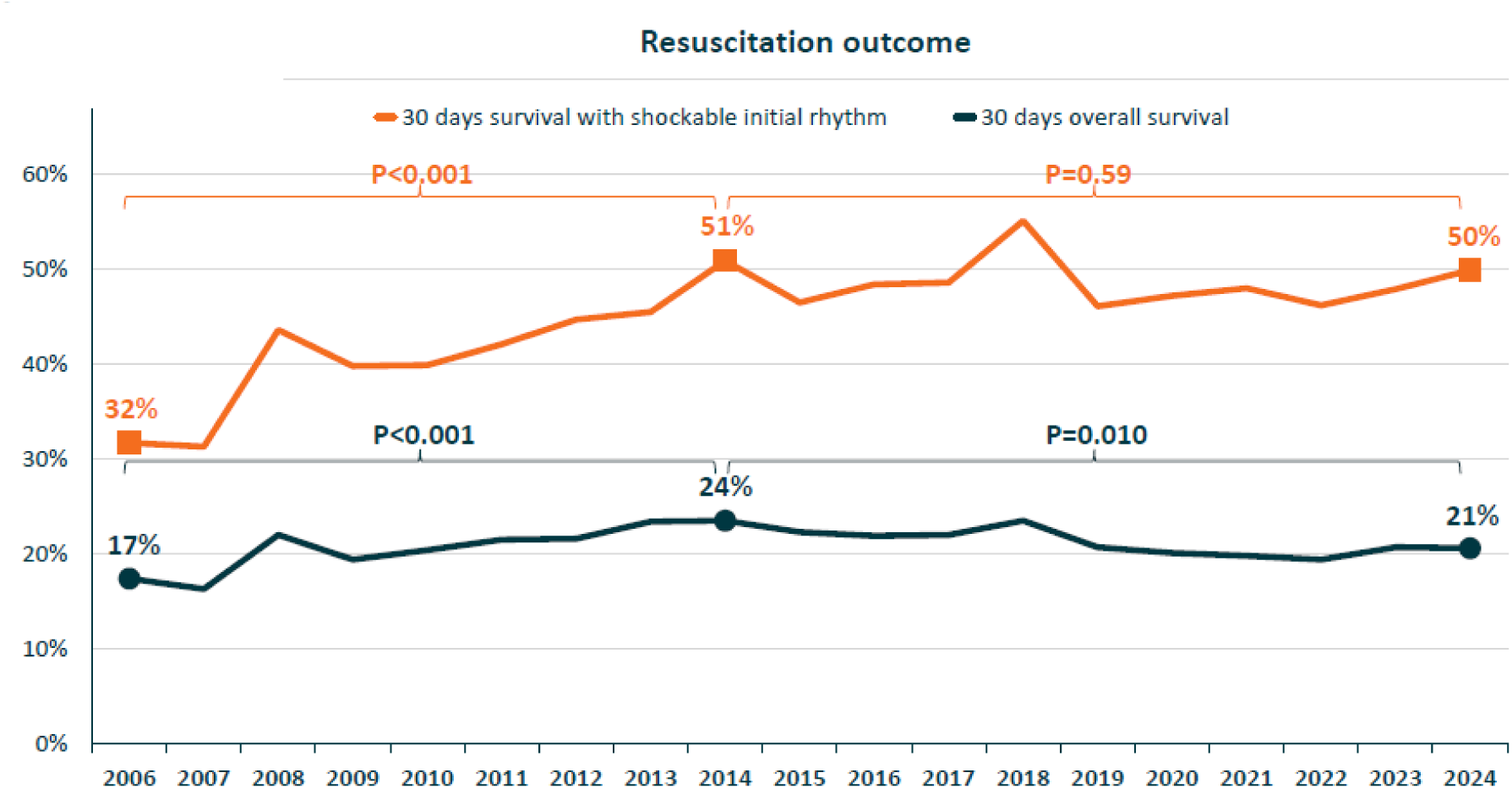
Trends in OHCA survival in the North-Holland province, as recorded in the ARREST Registry

The fundamental goal of CPR is to ensure an adequate supply of oxygenated blood to the brain and heart until spontaneous circulation is restored. While chest compressions are essential for maintaining cardiac output, effective ventilation is equally critical to maintain oxygenation. Without sufficient oxygen delivery, the chances of successful resuscitation and favorable neurological outcomes are severely diminished.^3, 4^ This is underscored by the fact that more than 50% of patients who survive to be admitted to the Intensive Care Unit ultimately succumb to extensive hypoxic brain injury.^5, 6^

Despite its vital role in CPR, ventilation remains underexplored. Historically, ventilation was often viewed as secondary to chest compressions or even potentially harmful, resulting in limited study and a lack of robust clinical evidence on key ventilation parameters such as tidal volume, ventilation rate, fraction of inspired oxygen and PEEP.^7^

### 1.2 Preclinical and observational evidence

Recent years have seen increasing interest in the use of PEEP to improve oxygenation during CPR.^8, 9^ Repetitive chest compressions cause rapid alveolar collapse, impairing oxygenation as circulating blood fails to adequately contact oxygenated alveoli.^7^ PEEP has long been a mainstay in critical care to prevent alveolar collapse by maintaining positive pressure in the lungs during expiration.

Previously, the use of PEEP during CPR was discouraged due to unsupported theoretical concerns about its potential negative effects, such as diminished venous return and cardiac output. Recent animal studies, however, demonstrate that PEEP can enhance oxygenation and oxygen delivery through alveolar recruitment, without detrimental effects on blood pressure.^10–12^ Nevertheless, levels above 10 cm H_2_O may decrease cardiac output to the extent that net oxygen delivery (DO₂) declines, despite preserved arterial pressure. A PEEP level of 8 cm H_2_O appears to strike an optimal balance—improving gas exchange without impairing cardiac output during CPR. By enhancing oxygen delivery, PEEP may help preserve cerebral oxygenation and limit hypoxic brain injury during CPR.

Observational studies have demonstrated an association between ventilation strategies incorporating PEEP and improved arterial oxygen content, as well as higher rates of ROSC, compared to ZEEP.^13, 14^ These findings highlight the potential clinical benefits of PEEP during CPR, supporting the rationale for further investigation in adequately powered trials.

Other strategies aimed at modulating intrathoracic pressure during CPR, such as the use of an inspiratory impedance threshold device to enhance negative intrathoracic pressure, have demonstrated promising physiological effects in preclinical models but have not translated into improved outcomes in a blinded randomized trial.^15^ This discrepancy highlights the challenges of translating physiological and animal data into real-world clinical practice, and underscores the need for robust randomized evidence when evaluating ventilation-related interventions.

### 1.3 Current practice and need for a randomized controlled trial

To date, clinical practice guidelines are based on expert opinion and do not provide clear recommendations regarding the use of PEEP during CPR.^16^ As a result, ventilation practices during CPR vary widely between centers; some continue to ventilate without PEEP (ZEEP), largely reflecting historical concerns regarding potential adverse haemodynamic effects, whereas others apply PEEP or use specifically designed CPR ventilation modes (CPV, CCSV) that incorporate PEEP, based on emerging physiological and experimental evidence suggesting improved alveolar recruitment and oxygenation during resuscitation (Figure 2).^17^

**Figure 2.**
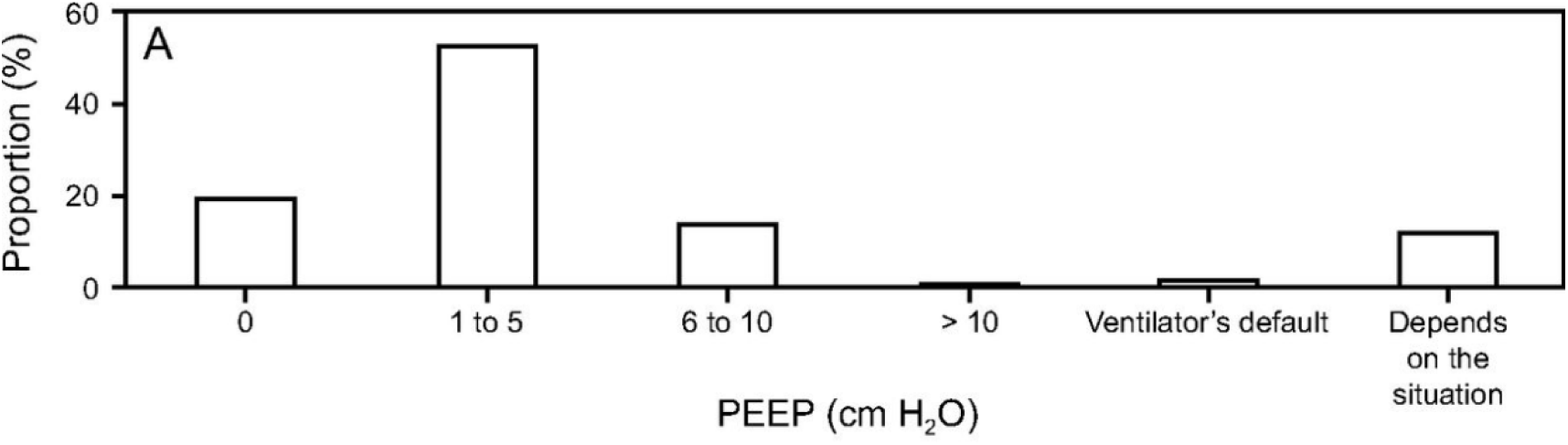
Results of an international survey on the use of PEEP during CPR, demonstrating substantial variability in reported ventilation practices and opinions.^17^

The International Liaison Committee on Resuscitation has explicitly recognized the paucity of randomized evidence on ventilation strategies during out-of-hospital cardiac arrest, identifying the absence of adequately powered randomized controlled trials—particularly those evaluating neurological outcomes—as a key knowledge gap.^18^ Although the potential PEEP to improve outcomes during CPR is biologically plausible, the reliance on limited observational data underscores the need for a large, well-designed randomized trial to provide definitive guidance on its use in clinical practice.

## 2. OBJECTIVES

We hypothesize that, during CPR with manual ventilation after placement of a SAD or ETT, use of a PEEP valve applying 8 cm H_2_O, compared with a sham valve resulting in ZEEP, improves neurological outcome at hospital discharge.

### 2.1 Primary Objective

The primary objective of this study is to assess if the intervention leads to an improved neurological outcome at discharge, as assessed by the utility-weighted modified Rankin Scale (UW-mRS).

Survival alone does not fully capture outcomes that are most meaningful to patients after OHCA. Interventions may increase the number of survivors, but some survivors may experience severe neurological impairment, which can greatly reduce quality of life. For example, the PARAMEDIC-2 trial showed that while epinephrine increased 30-day survival compared with placebo, it did not improve the rate of favorable neurological outcomes; as a larger proportion of survivors in the epinephrine group had severe neurological deficits.^19^ This underscores the importance of evaluating neurological function as a patient-centered outcome, rather than focusing solely on survival.

Although the mRS is widely dichotomized in cardiac arrest research, dichotomization discards important information contained in the rank ordering of outcomes. Ordinal analysis preserves the full range of outcome data, allowing for a more nuanced assessment of treatment effects across all health states.^20^ In addition, UW-mRS incorporates patient-centered preferences by weighting outcomes according to their impact on quality of life.^21^ In this study, we use a UW-mRS in which mRS-5 and mRS-6 are assigned the same weight, reflecting Dutch patient preferences regarding quality of life (table 1).^22^

**Table 1.**
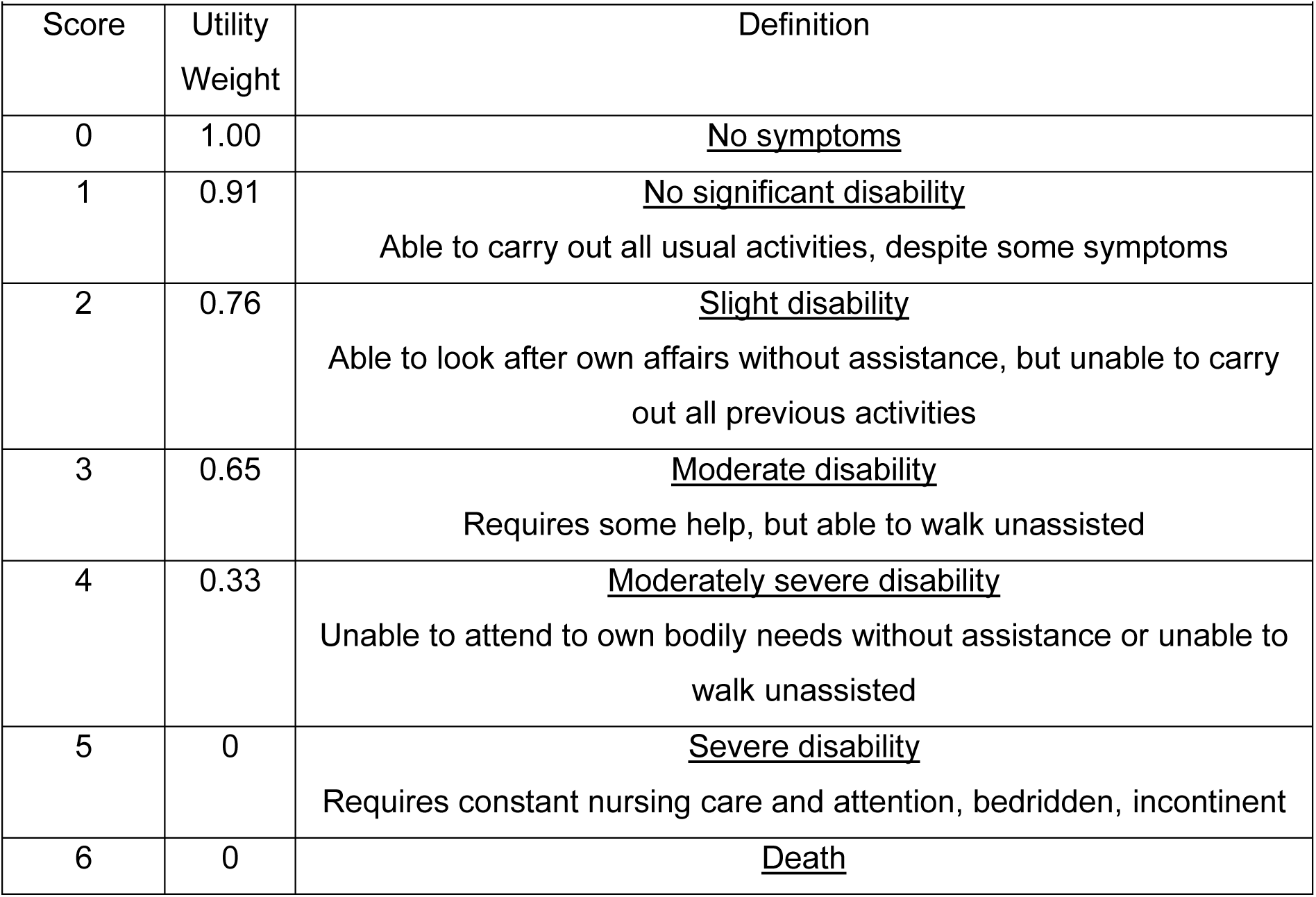
Utility-weighted modified Rankin Scale (UW-mRS)^22^.

Applying this approach to cardiac arrest research allows for a more comprehensive and patient-focused evaluation of neurological outcomes, rather than relying solely on survival or dichotomized functional status.

### 2.2 Secondary Objectives

The secondary objectives are of an exploratory nature and should be interpreted as preliminary findings. They aim to generate hypotheses for future studies rather than provide definitive conclusions.

- Clinical outcomes

○ To evaluate the effect of the intervention on ROSC.
○ To evaluate the effect of the intervention on dichotomized neurological favorable survival at discharge, as indicated by a score of 3 or less on the mRS.
○ To evaluate the effect of the intervention on 30-day survival.
○ To evaluate the effect of the intervention on health-related quality of life among survivors at 6 months after cardiac arrest, measured through the EQ-5D-5L questionnaire.
- Safety parameters

○ To evaluate the effect of the intervention on the occurrence of clinically significant pneumothorax, defined as the need for drainage through (needle) thoracostomy or chest tube.

### 2.3 Substudies

The following substudies focus on exploratory and mechanistic endpoints and are therefore not included in the primary publication but will be reported separately.

- To evaluate the effect of the intervention on pulmonary mechanics in a subset of patients, including peak pressures, effective PEEP, and lung compliance.
- To evaluate the effect of the intervention on airway opening index, a measure of intra-arrest intrathoracic airway closure.
- To evaluate whether the intervention affects thoracic impedance, reflecting changes in lung inflation during CPR.
- To evaluate the effect of the intervention on successful ventricular fibrillation termination, defined as the presence of a non-shockable rhythm 5 seconds after the shock.
- To evaluate the effect of the intervention on the intra-arrest blood gas parameters (pO₂, pCO₂, pH, base excess, bicarbonate, lactate) drawn in the hospital.
- To evaluate the effect of the intervention on atelectasis, defined as the fractional area of atelectasis on standardized CT scan if made as part of routine post-resuscitation care.
- To evaluate the effect of the intervention on the incidence of post-anoxic brain damage.

## 3. STUDY DESIGN

### 3.1 Overview

The REVIVE-PEEP trial is an investigator-initiated, pragmatic, registry-based, multicenter, parallel-group, triple-blind randomized controlled superiority clinical trial in the ARREST registry designed to evaluate the clinical effectiveness of adding 8 cm H_2_O PEEP to the standard advanced life support (ALS) protocol in patients with OHCA who have received a SAD or ETT.

This trial will be conducted within the ambulance services of the North-Holland and Flevoland provinces. In total, 3,200 patients will be enrolled. The sample size is based on a clinically meaningful 4% absolute reduction in mortality, reflected by an increase in favorable neurological outcomes within the modified Intention-To-Treat (mITT) population as assessed by the UW-mRS. The underlying assumptions and scenario analyses informing this calculation are described in Section 4.4.2.

### 3.2 Intercurrent events and estimand definition

In this trial, the intervention can only be delivered to patients who undergo advanced airway management, as patients receiving only bag-valve-mask ventilation cannot physiologically benefit from the intervention (see section 6.5). Therefore, initiation of advanced airway placement is considered a post-randomization intercurrent event, rather than an inclusion criterion, as randomization occurs at the start of CPR using pre-randomized kits, while airway management typically follows initial bag-valve-mask ventilation. Importantly, the assigned study valve (PEEP or sham) remains attached to the resuscitation bag throughout care, and manual ventilation is continued even after placement of an advanced airway (i.e. manual ventilation is not replaced by sustained mechanical ventilation).

The treatment effect of interest is defined by a principal stratum estimand, targeting the effect of PEEP in the theoretical subpopulation of patients who would require advanced airway management, regardless of treatment allocation. This principal stratum is not directly observable and reflects the estimand (i.e., the target of estimation), rather than a specific subset of patients.

The primary analysis will be performed in the modified intention-to-treat (mITT) analysis subset, consisting of patients in whom advanced airway management is actually performed and sustained mechanical ventilation is not used. This subset-based analysis serves as an estimator for the principal stratum estimand but should be interpreted with caution, as subsetting on observed intercurrent events does not, in general, correspond exactly to the principal stratum.

The mITT analysis provides an unbiased estimate of the principal stratum estimand under two key assumptions: (1) the allocation does not influence airway placement (i.e., there are ‘intervention initiators’ or ‘control initiators’), (2) advanced airway placement is clearly defined and measurable.^23^

In the REVIVE-PEEP trial, these assumptions are plausible. First, decisions regarding advanced airway placement are determined by EMS protocols, the provider’s airway experience, and patient need, rather than treatment allocation, which is blinded. In other words, being randomized to the PEEP or sham arm is highly unlikely to influence whether a patient receives advanced airway management. Second, these decisions are consistently documented in the ambulance records, ensuring transparency and reliable data.

### 3.3 Setting

In the Netherlands, two ambulances respond to presumed OHCA cases, both staffed by a driver and an ALS trained ambulance nurse, skilled in manual defibrillation, advanced airway management, drug administration, the use of automated chest compressions devices and vascular access. Ambulance nurses provide prehospital care according to national EMS guidelines. The local resuscitation protocols are based on Dutch and European Resuscitation Council guidelines.^24^ The ambulance nurses are allowed to terminate resuscitation on scene, based on established criteria.

### 3.4 Intervention

During the study, ventilation will be performed using a standard disposable resuscitator bag (SPUR II, AMBU A/S, Ballerup, Denmark) with a pre-assembled blinded PEEP or ZEEP/sham valve, provided as a single resuscitator kit. The same resuscitator bag will be used throughout the entire resuscitation attempt, both for bag-valve-mask (BVM) ventilation and for ventilation after airway management.

### 3.5 Blinding

The study is triple-blinded, ensuring that participants, ambulance staff, and researchers are unaware of group assignment. The intervention valve will provide 8 cm H_2_O of PEEP, while the sham valve in the control group will mimic the intervention valve in appearance but remain inactive, delivering ZEEP. Both valves will be identical in size, shape, and external markings. The triple-blind design reduces the potential for bias, though some minimal impact from subtle cues (e.g., slight acoustic differences between valves) cannot be entirely excluded. Outcome assessment and statistical analysis will also be carried out in a blinded manner.

For the interim analyses, a randomization list with group assignments will be shared with the independent statistician of the Data Safety Monitoring Board (DSMB). The randomization list, generated by Castor Electronic Data Capture system (Castor, version 2019), will be exported at the start of the trial by Research Data Management as a single list for all prerandomized randomization IDs that will be assigned to the CPR kits. Solely the independent statistician and medical technical department will have access to the randomization list to ensure the integrity and confidentiality of the blinding process.

### 3.6 Allocation

Batches of twelve pre-assembled resuscitator bags and PEEP valves—containing a 1:1 ratio of intervention and control devices in randomly permutated blocks of size 2 —will be prepared and sealed into individual bags by the medical technical department according to the randomization list. Each device will be clearly labeled as a study product, including the trial name, a unique trial ID, and a study contact phone number.

Allocation concealment is ensured by blinding of the ambulance personnel: during restocking, they will randomly select a device from the available stock, guaranteeing unbiased allocation for each case. After each CPR attempt, the ambulance team will restock, with the next CPR attempt typically not occurring during the same shift. Additionally, ambulance vehicles are randomly assigned to available staff, further ensuring that no influence from prior device selection affects subsequent allocation. In all resuscitation attempts, two ambulances respond, and the CPR kit from the first-arriving unit is used, consistent with clinical practice. The local investigator will oversee inventory management and setup at each ambulance post. Any issues or deviations will be reported to the coordinating investigator to ensure protocol adherence and integrity of the randomization process.

The trial randomization ID will be a mandatory field in the ambulance run report to verify the allocation. It will be included on a tag that can easily be torn off and stored during the CPR attempt, as well as on the product itself. If the patient is transported to the hospital, the trial ID will also be included in the hospital documentation, where feasible.

### 3.7 Procedures

All resuscitation attempts initiated by EMS will be included, irrespective of final eligibility, to preserve the pragmatic design. Patients will be considered randomized once ventilation with the study device is commenced during CPR. In the case of sustained ROSC (>20 minutes), ambulance personnel may remove the PEEP valve at their discretion if deemed clinically necessary.

The vast majority of data required for the study is routinely collected within the ARREST registry. The only additional tasks for ambulance staff are recording the trial identification number in the case report form and noting any reasons for patient exclusion or deviations from the protocol. To further explore physiological endpoints and validate treatment allocation, ventilation metrics will be collected for a subset of patients in a blinded manner, thereby preventing interference with study behavior.

### 3.8 Stakeholder involvement

Ambulance personnel have been involved as stakeholders from the development phase of this study to ensure the operational feasibility of the protocol in daily practice; this involvement is documented in the study team’s organogram. This input contributed to, among other things, the decision to randomize before rather than after advanced airway management (see §4.2), and to minimizing study-specific tasks for ambulance personnel during resuscitation (see §8.3). Prior to the start of inclusions, ambulance personnel will additionally be informed of the study procedures through an instructional video (see §6.9).

The patient perspective has been incorporated both in the design of the study and in the informed consent procedure, through collaboration with the patient association of the Dutch Heart Foundation. This collaboration included review, discussion, and approval of the patient information letter and consent procedure (see §11.3).

Ambu A/S, as the manufacturer of the investigational devices, has been involved as a partner throughout the development of the study valves, which are purpose-built for this clinical investigation.

## 4. STUDY POPULATION

### 4.1 Population (base)

For this trial, we have selected regional ambulance services where data is already being prospectively collected as part of the existing ARREST registry. This approach allows us to leverage the established infrastructure and robust data collection processes already in place, ensuring efficiency and consistency while maximizing the trial’s feasibility and reliability.

The EMS regions Amsterdam-Amstelland/Zaanstreek and Noord-Holland Noord collectively cover a population of approximately two million people, with historical data from 2023/2024 indicating 880 eligible OHCA cases per year for the mITT analysis. To accelerate patient inclusion, the study will utilize the ongoing incorporation of the Gooien Vechtstreek and Flevoland regional ambulance services into the registry. This will increase the covered population to approximately three million people, which is expected to raise the number of eligible cases in the mITT subset for the primary analysis by roughly 50% - to around 1,250 per year.

Assuming an ideal inclusion rate, the target sample size is expected to be reached within 2 years. To account for potential delays or slower-than-anticipated inclusion—a common issue in multicenter trials—a half-year margin has been added by extending the inclusion period to 2.5 years.

### 4.2 Inclusion criteria

All adult patients with OHCA who receive an advanced airway will be eligible for the primary analysis, regardless of initial rhythm, witness status, or whether CPR before EMS arrival was provided. To minimize workload for ambulance personnel and enhance data reliability, randomization will occur prior to advanced airway management. Based on historical data, we anticipate that approximately 25% of randomized patients will not be eligible for the primary analysis (see section 6.5).

Cardiac arrest is defined as unconsciousness, abnormal breathing, and loss of pulses requiring chest compressions and/or defibrillation. If resuscitation is not attempted by ambulance services, even when CPR was initiated by lay responders, due to the patient being obviously dead (i.e., rigor mortis, maceration, or putrefaction), the patient will not be included in the screening list.

### 4.3 Exclusion criteria

Any potential subject who meets the following criteria will be excluded from the study:

- Suspected traumatic cardiac arrest
- Suspected cardiac arrest after drowning
- Continuous use of mechanical ventilation

Traumatic OHCA (e.g. due to blunt or penetrating trauma) and cardiac arrest following drowning will be excluded due to their distinct pathophysiology. In contrast, arrests resulting from hanging, strangulation, or foreign body airway obstruction will be included. The trial will not exclude females of childbearing age or those with known or suspected pregnancy.

Short-term mechanical ventilation used to facilitate patient transport (e.g., during fire department hoisting) will be allowed, provided it is not initiated directly after airway management and used only during transport, without being continued beyond the immediate transport period. Participation in other clinical trials will not be generally restricted but will be assessed individually on a case-by-case basis.

### 4.4 Sample size calculation

The sample size is based on the primary endpoint of neurological outcome at hospital discharge within the mITT subset. We defined an absolute risk difference of 4% in survival as minimally clinically important and considered two scenarios for the neurological outcomes of these additional survivors.

#### 4.4.1 Minimally clinically important difference in survival

The study defines an absolute reduction of 4% of patients with mRS-6 (death) at discharge as the minimally clinically important difference between the control and intervention groups, based on the rationale that an intra-cardiac arrest intervention should lead to a significant improvement in survival in order to justify a change in clinical practice guidelines.^25^ Historical data from the 2023 ARREST registry indicate a 30-day survival rate of 12% in the target population, which is lower than average survival due to resuscitation time bias, as advanced airway management typically occurs during prolonged CPR.

#### 4.4.2 Scenarios for distribution of neurological outcomes

We considered two scenarios for the neurological outcomes of these additional 4% survivors: a Favorable scenario and an Unfavorable scenario. Both scenarios are explained graphically in Figure 3.

**Figure 3.**
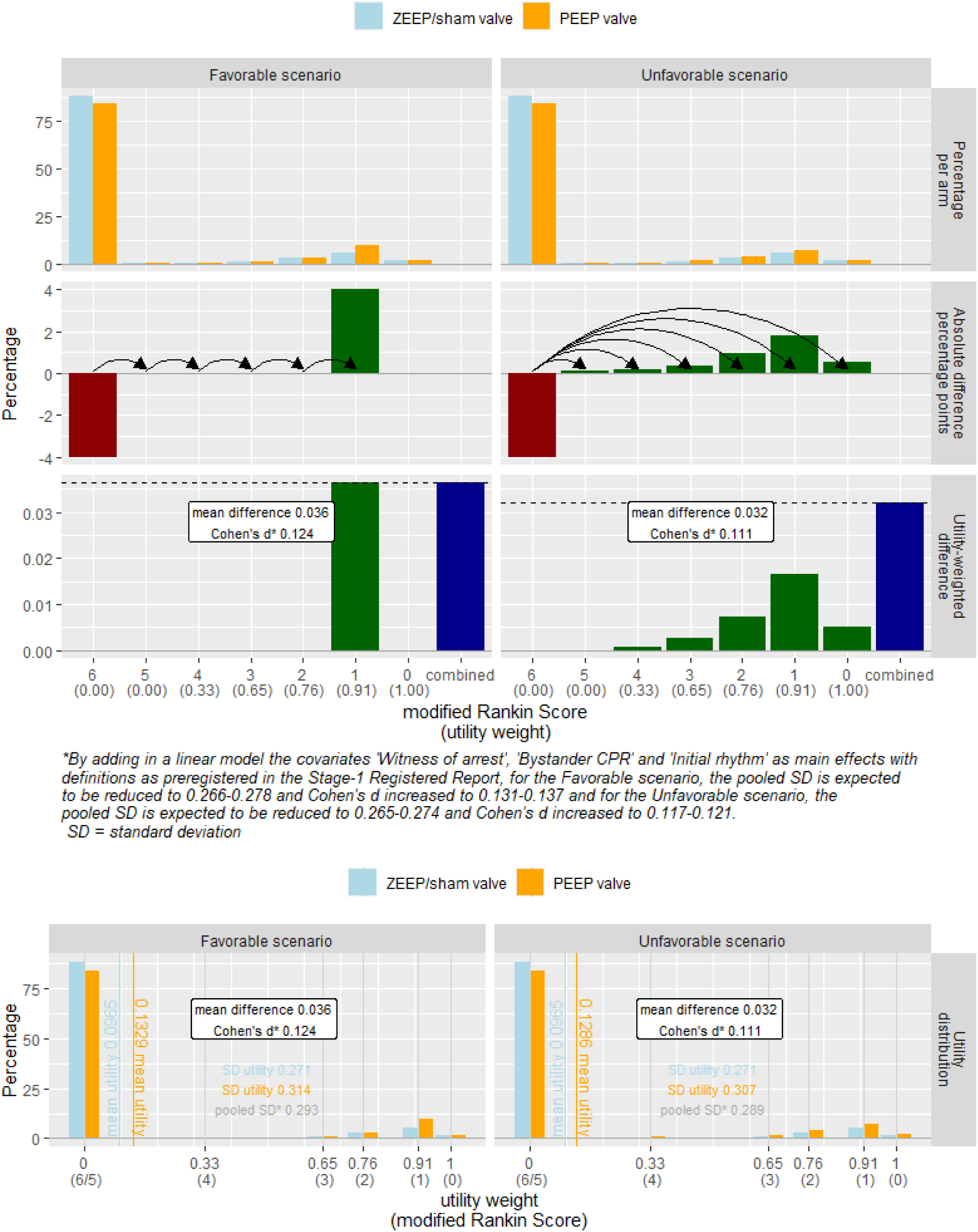
Graphical illustration of neurological outcome distributions

In the Favorable scenario, 4% of the total patients who would have died in the ZEEP/sham valve group now survive with mRS-5, while the same number who would have survived with mRS-5 improve to mRS-4, continuing similarly up to mRS-1. Given the very low likelihood of survival with mRS-0 in the ARREST database (<1%), additional survivors are assigned mRS-1. The Unfavorable scenario distributes the additional 4% survivors according to the observed survivor base rates in the ARREST database. Because some survive with mRS-5 (utility 0) and the differences between mRS-4 to mRS-2 are larger than between mRS-0 and mRS-1, the mean utility gain is smaller in this scenario than in the Favorable scenario. An Excel sheet is provided as supplementary material to illustrate Cohen’s d calculations for both scenarios.^26^

Cohen’s d quantifies the standardized mean difference in utility-weighted neurological outcomes between groups, and was used for sample size and power calculations. Figure 3 explains graphically how a Cohen’s d follows from the two scenarios. Sample size calculations were based on a two-sided 5% alpha and a Cohen’s d range of 0.117–0.137, corresponding to a minimum under the Unfavorable and a maximum in the Favorable scenario. These ranges are estimated from various subsets of existing ARREST data based on a regression model including the main effects ’Witness of arrest’, ‘CPR before arrival EMS’ (including EMS witnessed) and ’Initial rhythm’ that are prespecified for the primary analysis (see Section 10.1). With a sample size of 2,400 patients with data on neurological outcomes in the mITT analysis and six interim analyses conducted between 1,000 and 2,000 patients, utilizing an O’Brien-Fleming alpha-spending function, the trial achieves power ranging from 81% to 91% (see Appendix 1 for the results obtained from the software nQuery).

#### 4.4.3 Total sample size

Based on historical data, we anticipate a total 25% post-randomization exclusion rate for the mITT analysis. Therefore, the target enrollment is increased by a factor 1/0.75 to 1,600 participants per group, resulting in an estimated total sample size of 3,200 patients (2,400 * 1/0.75 = 3,200). The trial will be concluded once the required number of 2,400 participants with available neurological outcomes is included in the mITT analysis.

Given the pragmatic design, in which patients are automatically enrolled as part of routine practice, we expect high compliance with the study protocol. The target sample size for the mITT subset is 2,400 patients with available neurological outcomes, expected to be reached over 2.5 years. Figure 4 presents a preliminary CONSORT-style flowchart of planned study enrollment

**Figure 4.**
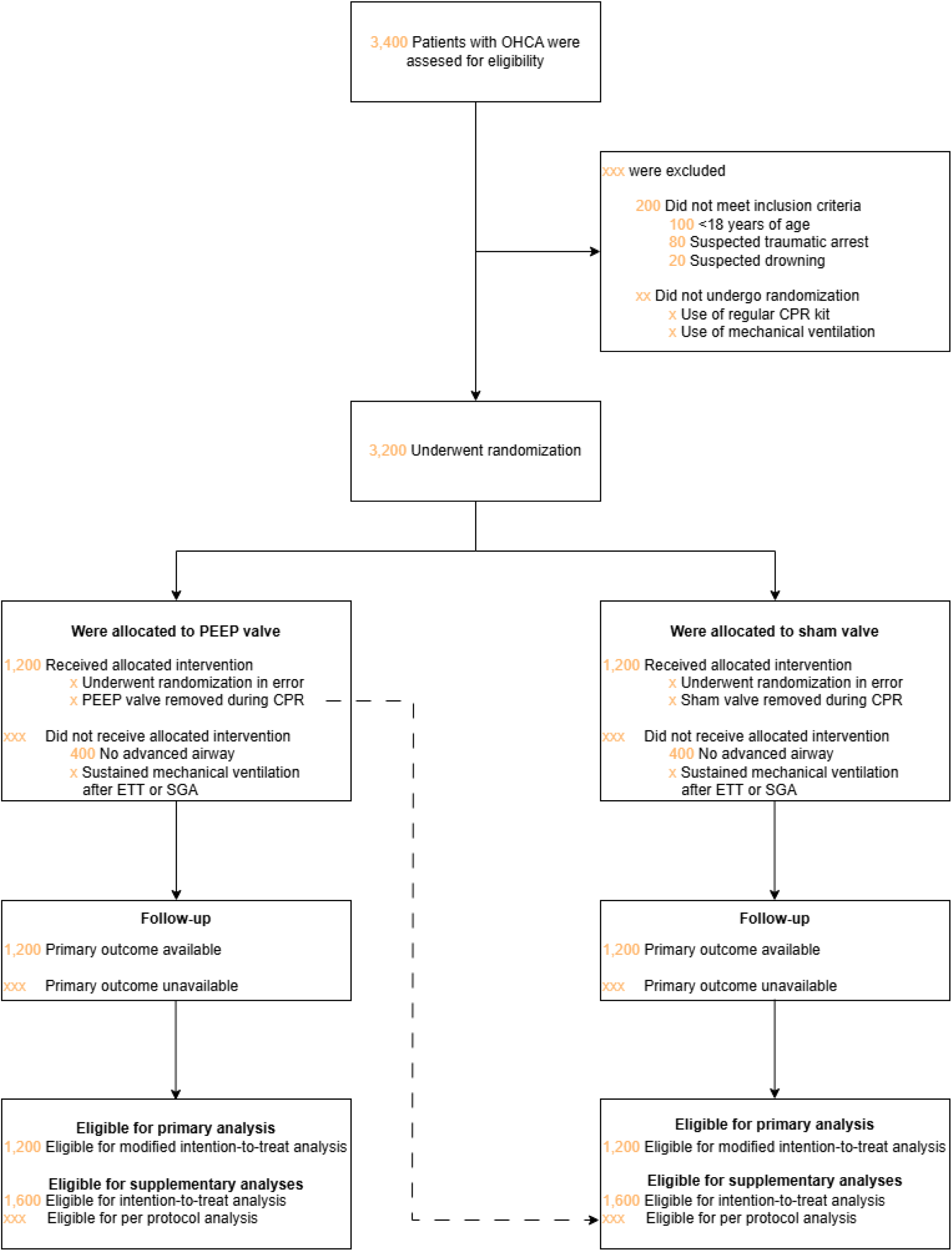
Planned Participant Enrollment Flowchart for REVIVE-PEEP trial

#### 4.4.4 Prospective meta-analysis

Part of the sample size plan is to recalculate the maximum sample size of patients at the 6^th^ interim analysis after inclusion of 2,000 patients to decide whether a new trial should be prepared to increase the sample size beyond this trial. This sample size re-estimation will be performed based on blinded group allocation, but with knowledge of the two possible efficacy analyses (coded as A/B, B/A). In case a new trial is started, it will be analyzed in a prospective meta-analysis with this trial.

## 5. TREATMENT OF SUBJECTS

### 5.1 Investigational product/treatment

In routine clinical practice, ambulance services provide manual ventilation, a critical component of which is the use of an adjustable PEEP valve. Adjustable PEEP valves are a standard part of the equipment in the ALS bag brought to the scene of critically ill or injured patients and are used both in and outside of CPR. Ambulance providers are extensively trained in the use of Ambu^®^ resuscitators and their associated valves to ensure optimal ventilation management during resuscitation efforts.

While resuscitators and adjustable PEEP valves are already market-approved and commonly used in clinical practice, this study will utilize a custom-made device specifically modified for blinding. The intervention involves a medical device designed to deliver a fixed level of 8 cm H_2_O PEEP during CPR. The placebo will consist of a blinded PEEP valve set at 0 cm H_2_O.

### 5.2 Use of co-intervention (if applicable)

All medications and interventions administered to subjects will follow the standard ALS protocol. The study does not interfere with or alter any aspects of this protocol. Any co-medications or additional interventions required as part of standard ALS care are permitted and will be managed according to established medical guidelines.

Airway management will adhere to local ambulance protocols aligned with European guidelines, which emphasize provider competence. Providers with a high tracheal intubation success rate will use an ETT, while others will use a SAD.

European guidelines recommend that when an advanced airway is secured, ventilation should be performed at a rate of ten breaths per minute while chest compressions continue uninterrupted. If an SAD results in gas leakage and inadequate ventilation, chest compressions should be paused to allow ventilation using a 30:2 compression-to-ventilation ratio. Each breath should be delivered over 1 second to achieve visible chest rise.

### 5.3 Inclusion in other studies

This trial will partly run in parallel with the LUCASVP trial (NL-009277) that started in January 2026 in a selective number of ten ambulance stations. In the LUCASVP study, mechanical chest compression devices are randomized to either pause for three or five seconds during synchronous ventilation pauses, with the primary outcome being the difference in the percentage of ventilation pauses in which two ventilations are given. The total planned inclusion is 692 patients, and it is a single blind (patients) study. Although the selection and randomization procedures of both studies are not expected to influence or interfere with each other, there is a potential for interaction in the outcomes.

Given that the REVIVE-PEEP trial is scheduled to start in September 2026, it is anticipated that approximately 5-10% of the REVIVE-PEEP trial population may overlap with LUCASVP participants. In the REVIVE-PEEP trial, patients in the mITT subset —those who have received airway management— are typically ventilated asynchronously, meaning that ventilations are delivered independently of the ongoing chest compressions. This differs from synchronous ventilation, where two ventilations are given after every 30 compressions. Only in a minority of cases, such as when a SAD leaks, might patients effectively be included in both studies at the same time.

Additionally, the interventions studied in the LUCASVP trial fall within the European Resuscitation Council guidelines norm for ventilation pauses. In clinical practice, both 3-second and 5-second pauses are used in different regions; for example, the Noord-Holland Noord protocol typically uses a 5-second pause, while the Amsterdam area typically uses 3 seconds. Current evidence does not support an effect of pause duration on clinical outcomes – moreover, assessing clinical outcomes is not the primary goal of the LUCASVP study.

REVIVE-PEEP is a pragmatic trial, with the objective of assessing the real-world impact of applying PEEP within the current ALS protocol. It acknowledges that slight variations in local protocols are common and reflects the clinical reality of these variations. While there is a theoretical risk of outcome interaction, the primary outcome — utility-weighted neurological function (UW-mRS) — is unlikely to be affected. The interventions are different in nature, with no evidence suggesting that the combined exposure to both trials would lead to significant interaction in the outcomes.

We therefore believe that conducting the trials in parallel - with clearly delineated procedures and predefined risk mitigation strategies - offers a feasible, scientifically robust, and ethically sound approach.

### 5.4 Escape medication (if applicable)

The ALS bag carried to the scene will include standard adjustable (non-blinded) CE-marked PEEP valves for patient categories excluded from the study, such as drowning cases, but where PEEP is recommended according to standard practice. In the case of suspected trauma cardiac arrest, the ambulance nurse will remove the study PEEP valve prior to the use of the resuscitator.

Additionally, if there is a clinical concern for elevated ventilation pressures or thoracic hyperinflation, for example in patients with severe asthma, the PEEP valve may also be removed at the clinician’s discretion.

## 6. INVESTIGATIONAL PRODUCT

### 6.1 Name and description of investigational product(s)

Ambu Disposable PEEP 00 Valve and Ambu Disposable PEEP 08 Valve Ambu PEEP valves have been widely used in clinical practice for over fifty years.^27^ Their CE certification and extensive use across healthcare settings demonstrate compliance with European health, safety, and environmental standards.

The standard valves have been modified to facilitate blinding and support the randomized study design. These valves are not intended as novel clinical products but serve solely as tools to enable the controlled investigation of PEEP levels. Although we have adapted a thoroughly tested product, Ambu A/S conducted comprehensive due diligence regarding its function, safety, and risk assessment.

An investigator’s brochure is available to provide detailed information about the investigational devices. Every custom-made PEEP valve was bench-tested before clinical use, and the reliability testing is documented and summarized in the Verification Summary Report.

### 6.2 Summary of findings from non-clinical studies

The external validity of non-clinical studies is limited due to anatomical and (patho)physiological differences between animals and humans. Where ventilation in animal studies is controlled, it may not fully replicate the complexity of real-world resuscitation efforts, where lung inflation occurs infrequently with bag-valve-mask ventilation during 30:2 CPR.^28^ Furthermore, manually delivered ventilation differs between patients in terms of tidal volumes and rates. The lack of consistent ventilation during resuscitation efforts in real-world scenarios complicates the translation of experimental study results to clinical practice.

However, the concepts explored in these studies provide valuable insights into the potential effects of PEEP during CPR. The following summarizes the key findings from the non-clinical studies:

- A widely cited trial demonstrated that hyperventilation with excessively high tidal volume and a ventilation rate of 30/min significantly increased mean inspiratory pressure, leading to reduced coronary perfusion pressure and lower survival.^29^ Although the tidal volume used was far above clinical norms the study raised lasting concerns about the hemodynamic consequences of ventilation. In contrast, a subsequent study using a tidal volume of 18 mL/kg (still three times higher than recommended) and a ventilation rate of 33/min did not observe significant hemodynamic compromise, suggesting that the extremes found in the initial study are not clinically relevant.^30^
- Voelckel et al. (2001) investigated the effects of adding PEEP during active compression–decompression CPR combined with an inspiratory impedance threshold valve in a porcine cardiac arrest model. After 8 minutes of CPR, animals were randomized to continued intermittent positive pressure ventilation alone or with stepwise increasing PEEP levels (2.5–10 cm H_2_O). In the IPPV-only group, arterial oxygenation progressively declined, whereas in the PEEP group oxygenation significantly improved with increasing PEEP. In addition, PEEP increased the diastolic aortic–left ventricular pressure gradient as well as systolic aortic pressure, consistent with enhanced indirect myocardial compression. Coronary perfusion pressure, defined as the diastolic aortic–right atrial pressure gradient, was not adversely affected by PEEP. Notably, airway pressure changes induced by chest compressions were greater in the presence of PEEP, suggesting improved transmission of intrathoracic pressure to the airway opening, which may contribute to increased blood flow during CPR.
- Levenbrown et al. (2020) performed a randomized, controlled animal study where CPR was performed using a mechanical chest compression device, with pigs ventilated at a tidal volume of 8 mL/kg and varying ventilation rates, and randomized PEEP levels of 0, 5, 10, 15, and 20 cm H_2_O for 9 minutes.^12^ Cardiac output and PaO₂ were measured to calculate oxygen delivery for each PEEP level. The results indicated that PEEP levels of 0–5 cm H_2_O optimized oxygen delivery and cardiac output, with PEEP 5 providing slightly higher oxygen delivery and a statistically insignificant lower cardiac output compared to PEEP 0. Although PEEP levels up to 20 cm H_2_O were shown to increase oxygen content in the blood, increasing PEEP to levels ≥10 cm H_2_O reduced cardiac output, with a marginal decrease of 0.3 L/min when comparing PEEP of 0 with 20 cm H_2_O. This decrease in cardiac output with PEEP levels of more than 10 cm H_2_O ultimately leads to the negative effects outweighing the benefits from increased oxygen levels, resulting in a decreased DO₂ in this porcine model.
- Renz et al. (2022) performed a prospective, randomized, controlled animal trial to investigate the effects of three different levels of PEEP (0, 8.2, and 16.3 cm H_2_O) during prolonged CPR (30 min) in pigs, both in both standard tidal volumes (9-10 mL/kg) with a ventilation rate of 10/min and ultra-low tidal volumes.^10^ The study found that higher PEEP levels, particularly at moderate levels (8.2 cm H_2_O), enhanced lung tissue recruitment, optimized gas exchange, and improved oxygenation. Additionally, driving pressures were reduced, and ventilation–perfusion ratios were improved. Importantly, no significant changes in mean arterial pressure were observed, indicating that higher PEEP levels can enhance oxygenation without further compromising hemodynamics.
- Kopra et al. (2024) randomized intubated pigs to receive either asynchronous ventilations (continuous chest compressions) or synchronous ventilations (30:2 compression-to-ventilation ratio), both with a PEEP of 10 cm H2O for 35 minutes.^11^ The study demonstrated that applying PEEP is feasible during both synchronous and asynchronous CPR, with similar outcomes observed in gas exchange and vital signs. Notably, PaO₂ levels were significantly improved compared to a historical cohort without PEEP. While arterial blood gases, lactate, and hemodynamic markers showed no significant differences between groups, the asynchronous protocol was associated with a higher incidence of mild pneumothoraces on post-mortem examination. These findings support the use of PEEP to enhance oxygenation and ventilation during prolonged CPR, regardless of ventilation strategy, though continuous chest compressions may carry an increased risk of pneumothorax.

### 6.3 Summary of findings from clinical studies

PEEP is a cornerstone of ventilation and is used in virtually every ventilated patient to prevent alveolar collapse and improve gas exchange. Emerging clinical evidence suggests that PEEP may offer meaningful benefits during CPR:

- Grieco et al. (2019) demonstrated that the size of chest compression–induced oscillations in the expired CO₂ signal can serve as an indicator of lung expansion during CPR.^8^ In a three-part study involving OHCA patients, a bench model, and cadavers, they found that increasing PEEP helps reopen the lungs, improves the transmission of chest compression force through the airway, and enhances ventilation. Importantly, their findings suggest that applying the lowest PEEP necessary to maintain airway patency is sufficient to improve ventilation without increasing intrathoracic pressure or causing adverse hemodynamic effects.
- Hernández-Tejedor et al. (2023) conducted a non-randomized prospective study involving 150 patients who were ventilated either manually or mechanically. The study found an association between better ventilatory status in arterial blood gases with mechanical ventilation and 5 cm H_2_O of PEEP compared to manual ventilation. However, since ventilation parameters were not measured in the manual ventilation group, it remains unclear what factors are driving the observed differences in outcomes.^13^
- Van Wulpen et al. (2024) investigated the pressure needed to reopen the lungs during out-of-hospital cardiac arrest (OHCA) using pressure–volume curves. By connecting pressure and flow sensors to the endotracheal tube, they performed low-flow inflations during CPR. This allowed the identification of the lower inflection point on the curve—the pressure at which the intrathoracic airways begin to reopen. This point is clinically important, as pressures below it lead to airway collapse, limiting effective ventilation and oxygenation. The study found evidence of intrathoracic airway closure in all ten patients, with a median reopening pressure of 5.56 cm H_2_O (IQR 4.80–8.23 cm H_2_O). These findings suggest that PEEP should ideally be set just above this threshold to maintain airway patency during CPR.
- Malinverni et al. (2024) conducted a retrospective analysis to examine the association between mechanical ventilation, including 5 cm H_2_O of PEEP, and manual ventilation in 2,566 OHCA patients. The use of mechanical ventilation with PEEP was associated with a significantly higher probability of ROSC, both in unadjusted (odds ratio: 1.28) and adjusted analyses (adjusted odds ratio: 2.16). A major limitation of the study is its retrospective nature, which could introduce biases and limit the ability to establish causality.

In conclusion, these studies suggest that the physiological benefits of PEEP observed in routine clinical care also apply during CPR. PEEP enhances oxygen delivery through alveolar recruitment, even in the low-flow conditions during resuscitation. A PEEP level of 8 cm H_2_O appears to strike an optimal balance—sufficient to maintain airway patency and improve oxygenation, without compromising cardiac output.

Additionally, two small pilot randomized controlled trials are currently underway (NCT06836830 and NCT06939335), with planned sample sizes of approximately 600 and 132 participants, respectively. Although both investigate PEEP during cardiac arrest, they differ significantly in methodology, study design and sample size.^31^ While these studies will provide valuable insights, they are not expected to be sufficiently powered or methodologically comprehensive to independently inform guideline changes or allow definitive conclusions.^25^

### 6.4 Summary of known and potential risks and benefits

The longstanding belief that increased intrathoracic pressure during CPR reduces survival largely originates from a single, widely cited animal study, which used excessive ventilation settings not representative of clinical practice. Subsequent animal and clinical studies have contradicted these findings and instead provide evidence that PEEP levels below 10 cm H_2_O optimize oxygenation and ventilation without compromising cardiac output or hemodynamic stability. Preclinical and emerging clinical evidence supports that appropriately titrated PEEP can enhance alveolar recruitment and oxygen delivery during CPR, striking a balance that avoids detrimental cardiovascular effects.

Current guidelines do not raise concerns about PEEP’s effects on cardiac output but instead suggest that PEEP increases transthoracic impedance and should be “minimized during defibrillation” when possible.^24^ However, these recommendations lack support from clinical evidence. To provide context, even fully expanded lungs contribute less than 15% of electrical resistance in healthy volunteers and likely even less in patients undergoing CPR as a result of atelectasis.^32^ A PEEP level of 10 cm H_2_O increases electrical resistance by only about 3% in healthy volunteers.^33^ Moreover, modern biphasic defibrillators measure transthoracic impedance before defibrillation and adjust the delivered energy accordingly, rendering them largely unaffected by such small impedance changes. Clinically, defibrillation success rates are approximately 70-90%, and there is no evidence to suggest that PEEP adversely affects defibrillation effectiveness.

In summary, while theoretical risks remain a consideration, current evidence suggests that moderate PEEP use during CPR offers physiological benefits without the previously feared hemodynamic compromise. Further clinical research is needed to definitively clarify the risk–benefit profile of PEEP in this setting.

### 6.5 Description and justification of route of administration and dosage

To build up airway pressure, it is essential to prevent leakage, which requires an adequately sealed airway. Clinically, this can only be reliably achieved through advanced airway management, as a proper seal cannot be consistently maintained with BVM ventilation.

Therefore, the mITT subset includes only patients with advanced airway management and excludes those ventilated solely with BVM.

### 6.6 Dosages, dosage modifications and method of administration

The appropriate PEEP dose for this intervention was determined from both preclinical and clinical studies. The highest tolerable PEEP dose appears to be <10 cm H_2_O, as supported by preclinical studies showing hemodynamic detriment above this level. Clinical studies have identified the lower inflection point on the pressure-volume curve during CPR, suggesting that the optimal PEEP dose should be ≥8 cm H_2_O. A target dose of 8 cm H_2_O, with a ±1.5 cm H_2_O tolerance, provides an ideal balance between effectiveness and safety, remaining within the maximum tolerated range and below the threshold for adverse hemodynamic effects.

For the control group, an identically looking sham valve is used that is adapted to deliver 0 cm H_2_O of PEEP (i.e., ZEEP), with a tolerance of +0.3 cm H_2_O. This ensures the absence of clinically relevant PEEP while maintaining identical handling characteristics and appearance to the active intervention, thereby preserving blinding and minimizing performance bias.

After advanced airway management, asynchronous ventilation is primarily used. However, when leakage occurs with a SAD, current guidelines recommend switching to synchronous ventilation to improve the airway seal and ensure optimal pressure delivery. Studies have shown that PEEP provides benefits during both synchronous and asynchronous ventilation.

### 6.7 Preparation and labelling of Investigational Medicinal Product

AMBU will supply the SPUR II resuscitators and the study-specific PEEP valves as separate components. These will be stored under optimal conditions in AMBU’s storage facility at Schiphol, and transferred to the Amsterdam UMC in batches matched to the expected usage rate in the study. Products will be assembled by the medical technical service of Amsterdam

UMC and labeled according to the randomization list. A double-check will be performed during assembly following a four-eye principle to ensure accuracy and adherence to the randomization.

A distinct yellow color, not typically used for commercially available PEEP valves, will be applied to the study valves to clearly differentiate the investigational product from standard, commercially available devices (Figure 5). The packaging will also clearly state that these devices are only to be used for the REVIVE-PEEP trial. After assembly and labeling, the devices will be resealed in specially designed plastic bags to preserve integrity and ensure blinding for EMS personnel. The sealed, labeled devices will be stored under controlled conditions consistent with the manufacturer’s recommendations at the Amsterdam UMC and at designated logistical partners supporting the participating ambulance posts for short-term holding prior to deployment in the study.

**Figure 5.**
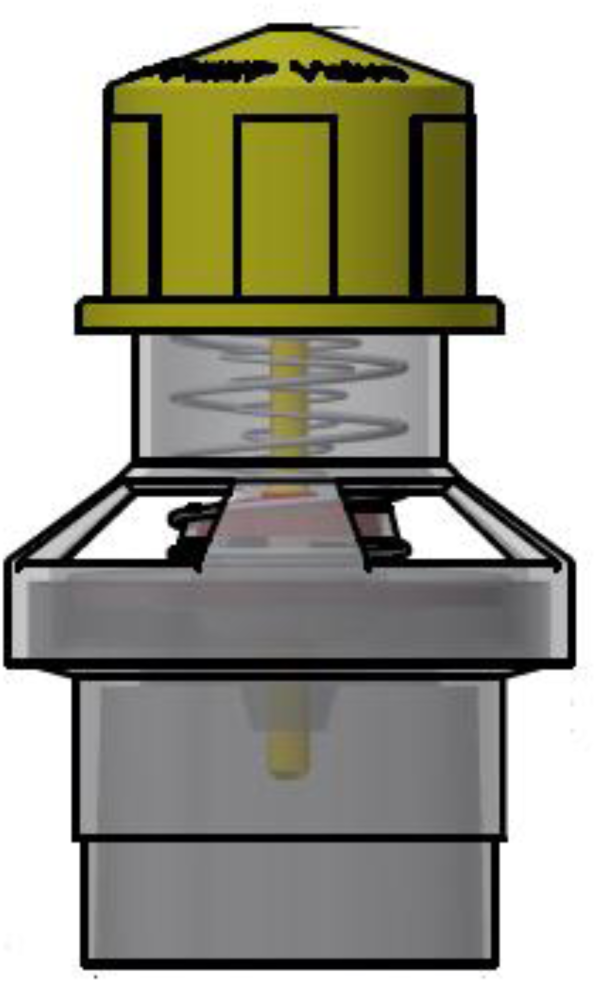
Design of the Study Devices

**Figure 6.**
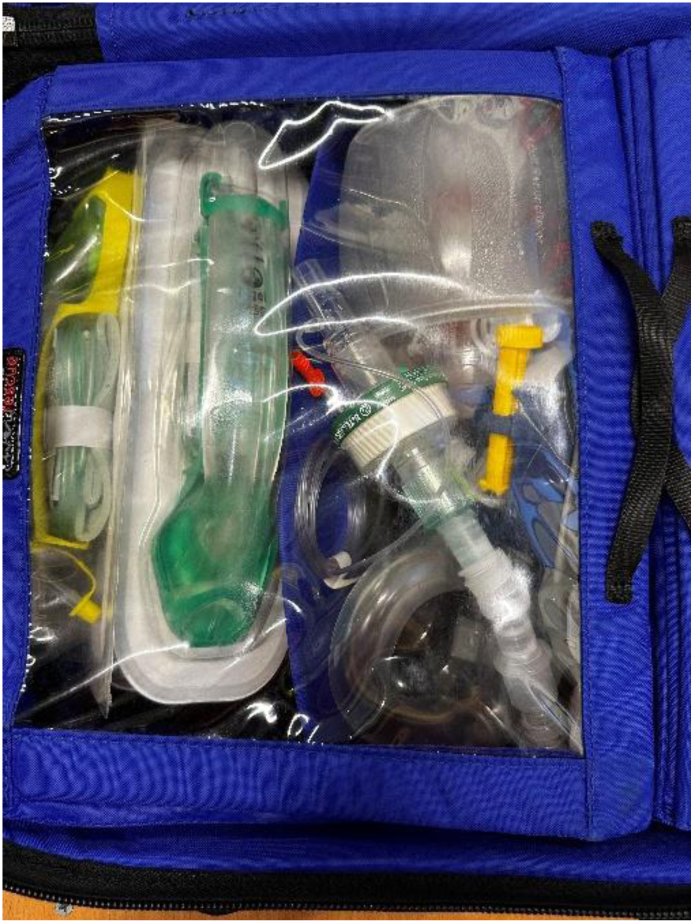
ALS bag with Ambu resuscitator

### 6.8 Device accountability

PEEP valves will be distributed from the central storage location according to the randomization list, which is numbered sequentially from lowest to highest. Products are allocated to sites based on the number of valves required at each ambulance post, ensuring supply matches anticipated patient volume. While a total of 7,000 study valves are produced in a one-off manufacturing process, each local post is expected to use only a few hundred per year. Centralized storage allows for gradual, demand-based distribution without overburdening local space and helps maintain consistency and adherence to the study protocol across all sites.

Study products will be stored in the ALS bag carried by EMS personnel during patient care on scene and are to be used exclusively for OHCA patients in accordance with the study protocol (figure 5). All EMS personnel will receive training on the correct use, handling, and documentation of the study devices. Any deviations from protocol use will be documented in patient records and reported according to study procedures to ensure compliance and accurate inventory tracking.

It is recognized that ambulance professionals must exercise clinical judgment in the field when responding to diverse patient presentations. While ambulance crews are encouraged to adhere to study indications and use alternative equipment when OHCA is deemed unlikely, operational and safety considerations may necessitate flexibility. In situations where immediate access to alternative equipment is not available, the use of the resuscitator bag without the study valve may be permitted to ensure timely patient care outside the context of cardiac arrest (e.g. intoxication, neurotrauma).

The PEEP valves and resuscitator bags are single-use disposable products and will be discarded after use. At the conclusion of the study, all unused study products remaining at participating sites will be retrieved to ensure accurate inventory records and minimize waste.

These will be transferred back to AMBU for disposal, with the exception of a reserve of 20 valves, which will be retained in storage for potential future reference, training, or other unforeseen study-related needs.

### 6.9 Training of Personnel on Investigational Product

Ambulance personnel are already familiar with the use of PEEP valves as part of their standard care protocol for OHCA patients. As such, there is no need for additional training on the basic use of the devices. However, study-specific information regarding the procedures, inclusion and exclusion criteria, and protocol adherence will be communicated to all involved personnel through a concise, approximately 5-minute instructional video developed in collaboration with the education department. This video can be integrated into regular CPR training sessions and will provide a standardized overview of the study procedures and correct application of the study devices. Additional communication will include newsletters and periodic reminders to reinforce key study information and ensure protocol adherence.

To further support the field personnel, local ambassadors will be selected at each ambulance post. These ambassadors will serve as on-site, direct points of contact for any questions or clarifications regarding the study protocol and investigational product. This ensures that staff have easy access to support and can address any concerns during the course of the study.

## 7. NON-INVESTIGATIONAL PRODUCT

In a subset of patients, respiratory monitoring devices will be used in fully blinded mode to record ventilation parameters during CPR. These devices will not influence clinical care or study procedures and will remain inaccessible to personnel involved in patient inclusion, data collection, or outcome assessment.

Devices must continuously measure airway pressure, airflow, and volumetric capnography, with a recommended sample rate of at least 25 Hz to capture dynamic changes during chest compressions. We aim to include this monitoring in a few hundred patients to provide sufficient physiological data for secondary analyses.

The data collected from these devices will provide valuable physiological insights for secondary analyses and will help verify protocol adherence as reported in the CRF. Specifically, pressure measurements will allow for the distinction between patients in the ZEEP and PEEP groups, confirming whether they received the assigned intervention as per the study protocol.

As these data effectively deblind the intervention, access to these data will be restricted and they will not be collected by, nor accessible to, the researchers involved in patient inclusion, data collection, trial coordination or outcome assessment.

## 8. METHODS

### 8.1 Study parameters/endpoints

Data for this study will be collected through the ARREST registry, which has continuously recorded all resuscitation attempts in the North Holland province since 2005. ARREST employs the Utstein OHCA Resuscitation Registry Template, a standardized framework for uniformly defining, collecting, and reporting cardiac arrest data, enabling consistent comparison of outcomes across studies, systems, and regions.^34^ This study will leverage ARREST’s established infrastructure, including its experienced team of data managers and PhD students, who ensure data quality and consistency through a detailed data dictionary aligned with Utstein standards.

#### 8.1.1 Main study parameter/endpoint

The primary objective of this study is to assess if the intervention leads to an improved neurological outcome assessed by the utility-weighted modified Rankin Scale (UW-mRS) at discharge, as discussed in section 2.1. Neurological outcome will be determined using information obtained from hospital medical records and the discharge summary.

#### 8.1.2 Secondary study parameters/endpoints

| Endpoint | Definition | Time point |
| --- | --- | --- |
| <u>Safety Endpoint</u> |  |  |
| Clinically significant pneumothorax | Needle thoracostomy or chest tube placement | During CPR or <1 hour of admission to hospital |
| <u>Clinical endpoints</u> |  |  |
| Any ROSC | Signs of circulation with no further use of chest compressions | During CPR attempt |
| 30-day survival | All-cause mortality | 30 days after CPR attempt |
| Neurologically favorable survival | mRS dichotomized as favorable (mRS 0-3) versus unfavorable (mRS 4-6) | At discharge |
| Quality of life | EQ-5D-5L questionnaire | 6 months ± 2 weeks after resuscitation attempt |

#### 8.1.3 Exploratory and mechanistic parameters/endpoints

| Endpoint | Definition | Time point |
| --- | --- | --- |
| Pulmonary mechanics | Average ventilatory parameters such as peak pressures, effective PEEP, and lung compliance. | During CPR after airway management |
| Intra-arrest blood gas parameters | pO<sub>2</sub>, pCO<sub>2</sub>, pH, bicarbonate, lactate | During CPR |
| Airway Opening Index | Calculated according to different proposed methods <sup>8, 35</sup> | Average values and trends over the course of the resuscitation attempt following airway management |
| Thoracic impedance | Defibrillator measured thoracic impedance during pre-shock pause | Average values and trends over the course of the resuscitation attempt following airway management |
| Fractional area of atelectasis | Automated segmentation of lung tissue in CT scans performed using an open-source artificial intelligence algorithm and manual retouches <sup>36, 37</sup> | Within 1 hour of hospital admission |
| Unsuccessful termination of VF | Presence of a shockable rhythm | 5 seconds after defibrillation |

#### 8.1.4 Other study parameters

The previously mentioned Utstein template encompasses a core dataset designed to facilitate comparison across clinical studies. It includes comprehensive and standardized information across several domains, such as:

- System-level data: cardiac arrests attended, resuscitation attempts, and reasons for no resuscitation (e.g., DNAR, obvious death, signs of life).
- Patient characteristics: age, sex, witnessed arrest, arrest location, CPR before arrival EMS and AED use, first registered rhythm, and presumed cause of arrest,
- Process measures: response time, time to first defibrillation, drugs administered, coronary angiography/STEMI status, prehospital airway management (BVM, SAD, ETT), time to airway placement, capnography waveform, use of mechanical CPR and ECPR, and inclusion and randomization in the LUCASVP trial.

An elaborative list, including definitions, data sources, and timepoints for each variable, is provided in Supplement 1.

### 8.2 Randomization, blinding and treatment allocation

The randomization list will be generated using Castor’s default system, employing randomly permuted blocks of size 2 in a 1:1 ratio for intervention and control groups to optimize for balance within the batches created. The medical technical department will receive the full randomization list from the department of research data management and assemble and label the study products according to this list. Batches of 12 products will be distributed to ambulance posts, where random restocking of devices will ensure patient-level randomization during routine use. A local research nurse will oversee inventory management and setup at each ambulance post. Any issues or deviations will be reported to the coordinating investigator to ensure protocol adherence and maintain the integrity of the randomization process.

In cases where breaking the randomization code is necessary—such as during serious adverse events or interim analyses—the independent DSMB statistician will be granted access to the randomization list. Unblinding procedures will be performed by the DSMB, and only authorized personnel, who are not involved in the daily conduct of the study, will have access to this information. This ensures that trial integrity is maintained to the greatest extent possible.

Patients, ambulance nurses, and researchers will remain blinded to intervention assignment until completion of data analysis. Study personnel will not have access to respiratory measurements, if obtained, during the conduct of the study.

### 8.3 Study procedures

Participants in this study will not undergo any additional procedures beyond those routinely performed as part of standard critical care, as all data collected originate from the ARREST registry. An exception applies to a subset of study participants in whom respiratory parameters are measured during CPR using a blinded, non-investigational device, as described in paragraph 7. These measurements are not part of the standard ARREST dataset but are included for specific research purposes.

The ARREST registry is based primarily on routinely collected clinical data during and after OHCA. This includes information from EMS and hospital records, as detailed in the ARREST research protocol and associated data collection instructions (“Onderzoeksprotocol ARREST 2.1”, section 5; “Werkinstructies ARR WI-01 Invoervelden ARREST database”; and “ARR WI-06 Ziekenhuisinformatie v3”).

mRS and scores are derived from hospital records and discharge letters. In addition, the EQ-5D-5L has been incorporated into the ARREST dataset to capture patient-reported outcomes and support more detailed neurological assessment at a later time point. Completion of the EQ-5D-5L during telephone follow-up is the only component that involves direct input from patients.

The additional workload for ambulance nurses is minimal. They perform resuscitation exactly as per standard protocols. The only study-specific actions required are a brief training on study inclusion and exclusion criteria and documenting the study number of the resuscitator in a mandatory field of their case report form. No additional procedures or deviations from routine care are necessary.

### 8.4 Withdrawal of individual subjects

Due to the nature of emergency research, a deferred consent procedure is used, as described in section 11.3. Subjects (or their legal representative) are notified of their participation in this study only after treatment and data collection have been completed. At that point, they have the opportunity to withdraw consent for the use of their data collected up to that time. Withdrawal can be done at any time, for any reason, and without any consequences for the subject.

### 8.5 Replacement of individual subjects after withdrawal

Consent is required for a subject’s data to be used in the primary analysis; where consent is declined or withdrawn, the corresponding data cannot be used and are not replaced on an individual basis. If the consent rate during the course of the study is substantially lower than anticipated — to the extent that it meaningfully threatens the trial’s statistical power — the sponsor/coordinating investigator will consult the DSMB and the accredited METC to determine an appropriate course of action.

### 8.6 Follow-up of subjects withdrawn from treatment

As the intervention occurs during the arrest, followed by ICU admission, the most critical period for monitoring adverse events is during their stay in the ICU, where participants are closely observed. Given that no realistic repercussions are expected after discharge from the ICU, no additional follow-up is required to monitor for adverse events related to study participation.

### 8.7 Premature termination of the study

The study will only be stopped prematurely if required by the DSMB/METC, as outlined in section 9.1.

## 9. SAFETY REPORTING

### 9.1 Temporary halt for reasons of subject safety

In accordance with section 10, subsection 4, of the WMO, the sponsor will suspend the study if there is sufficient ground that continuation of the study will jeopardize the subject’s health or safety. The sponsor will notify the accredited METC without undue delay of a temporary halt including the reason for such an action. The study will be suspended pending a further positive decision by the accredited METC. The investigator will take care that all subjects are kept informed.

### 9.2 AEs, SAEs and SUSARs

Historical data show a survival rate of 12% in our control group, indicating that serious adverse events (death, hospitalization, persistent disability or incapacity) unrelated to the trial intervention are expected in the vast majority of participants due to the underlying characteristics of the patient population.

Based on current literature, the only specific adverse event potentially attributable to the intervention is an increase in the incidence of mild pneumothorax. This condition can be managed conservatively and is therefore considered to be outweighed by the potential benefit in neurologically intact survival. A safety parameter for clinically significant pneumothorax is included among the secondary outcome measures and will be reported to both the DSMB and METC.

Because AEs, SAEs and SUSARs are expected to occur in >88% of patients, these events will not be reported individually. Instead, a line listing of the primary endpoint will be prepared by the DSMB and provided at each interim analysis, serving as a safety report to monitor both treatment strategies.

### 9.3 Data Safety Monitoring Board (DSMB)

The DSMB monitors efficacy and safety data at six interim analyses (see 10.6 for details), focusing on neurological outcomes at discharge, and considers secondary outcomes like ROSC and 30-day mortality only if they raise concerns about the primary outcome. Safety data on the occurrence of clinically significant pneumothorax, are also reviewed at six interims and ad hoc if needed, with secondary safety outcomes evaluated cautiously and only if relevant to the primary outcome. There are no formal stopping criteria for futility.

Additionally, the DSMB oversees overall trial conduct, including protocol compliance, recruitment, data quality, and completeness, as well as adherence to previous recommendations.

The DSMB’s recommendations will be communicated both to the study sponsor and the METC. If the sponsor chooses not to fully implement the DSMB’s advice, the sponsor must submit the recommendation to the reviewing METC, accompanied by a justification explaining why the advice (or parts of it) will not be followed.

## 10. STATISTICAL ANALYSIS

The study will adhere to the Consolidated Standards of Reporting Trials guidelines.^38^ In general categorical data (such as most patient characteristics) will be presented as counts and percentages. Continuous variables will be expressed as means (±SD) for normally distributed data (such as patient age), and as medians with interquartile ranges for non-normally distributed data (such as time until ambulance arrival).

The primary and secondary analyses will be performed in the mITT subset, defined as all randomized patients allocated to the study intervention who experienced the intercurrent event of advanced airway management without sustained mechanical ventilation.

### 10.1 Primary study parameter

The primary analysis will weigh the neurological outcomes on the mRS at discharge with the weights shown in Table 1 and adjust the estimate for the mean difference in utility by the known prognostic covariates ‘Witnessed Arrest’ (yes/unknown-no), ‘Bystander CPR’ (yes/unknown-no), and ‘Initial rhythm’ (shockable/unknown-non-shockable) as predefined main effects in a linear model.

Results will be reported as 95%-confidence intervals for the mean difference in utility based on a Gaussian approximation constructed at 1,000, 1,200, 1,400, 1,600, 1,800, 2,000 and 2,400 patients by margins of 3.280, 2.996, 2.766, 2.579, 2.427, 2.299 and 2.050 times the standard error respectively, shown as the upper efficacy bound in Appendix 1.

There are three possible primary conclusions:

I. The confidence interval for the mean difference in utility includes only positive values that indicate a benefit of PEEP. In this case, we will recommend to implement 8 cm H_2_O PEEP in CPR in adult patients suffering an OHCA not related to trauma or drowning, also in case of a small effect – given that cost and burden of implementation are low, while gain of each percentage point benefit is high, equaling hundreds of lives saved in The Netherlands and ten thousands worldwide.
II. Either the confidence interval for the mean difference in utility includes only negative values that indicate harm of PEEP, or we find safety concerns on the occurrence of clinically significant pneumothorax. In this case, we would recommend against routine implementation of manual PEEP at 8 cm H_2_O during CPR. These findings would not preclude further investigation of alternative PEEP levels in future studies.
III. We find no safety concerns and an inconclusive result with effects of benefit and harm both in the confidence interval. In this case, we recommend further studies to draw conclusions in a meta-analysis, while taking into account feasibility (e.g. value of information). In addition, we will explore the physiological factors behind the

primary conclusion based on key secondary outcomes included in the ARREST registry

### 10.2 Sensitivity and supplementary analyses

As a sensitivity analysis, the difference in mean utility weighted mRS scores will be analyzed without correction for the predefined covariates in a two-sample t-test with corresponding 95%-confidence interval, corrected for interim analysis analogous to the primary analysis.

For completeness, supplementary analyses for the primary endpoint will be performed using both intention-to-treat and per-protocol analyses. The intention-to-treat population is defined as all randomized OHCA patients allocated to the study intervention, irrespective of protocol adherence (valve removal, sustained mechanical ventilation) or airway management. The per-protocol population will consist of patients who received the study intervention as intended and underwent advanced airway management, with the patients with the valve removed in the sham/ZEEP group.

### 10.3 Secondary study parameters

Secondary outcomes will be compared between treatment groups using regression models correcting for the same main effects as in the primary analysis with appropriate outcome distributions: binomial for binary outcomes (e.g. ROSC, 30-day mortality, dichotomized mRS scores), Gaussian for continuous outcomes (quality of life)) and corresponding link functions (e.g., logit for binary outcomes, identity for continuous outcomes). Results will be presented as effect estimates with 95%-confidence intervals without correction for interim analyses and described in detail in figures and tables.

### 10.4 Other study parameters

Ventilation parameters (PEEP, maximum airway pressure, tidal volume, and ventilation rate) will be summarized descriptively for each treatment group. All differences will be reported with 95% confidence intervals assuming normality of means given large sample size. These intervals will not be corrected for interim analyses and covariates.

### 10.5 Subgroup analyses

#### 10.5.1 Key subgroup analyses

The primary outcome will be analyzed in the mITT subset according to airway modality (SAD versus ETT) and initial rhythm (shockable versus unknown/not shockable) as key subgroup analyses. Unlike other subgroup analyses, which are considered exploratory, this analysis is hypothesis-driven. We consider it a-priori plausible that treatment effect modification will occur, with a larger effect of PEEP expected in patients receiving ETT ventilation compared to those receiving SGA ventilation and larger effect expected in patients with a shockable rhythm.

Effect modification will be assessed for these two key subgroups in two separate regression models, both including all covariates from the primary analysis as well as the subgroup main effect and treatment-covariate interaction. A 5%-level one-sided test will be used to evaluate this interaction.

#### 10.5.2 Other subgroup analyses

Subgroup analyses will be performed to explore the following potential effect modifiers: sex, age, initial rhythm, bystander CPR, witnessed arrest, and ventilation mode after airway management (synchronous versus asynchronous).

Subgroup analyses will be conducted in the mITT subset for the primary outcome and the key clinical secondary outcomes (ROSC, 30-day survival, dichotomized favorable neurological survival and quality of life) using regression models with treatment × subgroup interaction terms to explore potential effect modification. They are considered exploratory.

### 10.6 Interim analysis

In total, six interim analyses are planned. The 95%-confidence intervals for the mean difference in utility based on a Gaussian approximation constructed at 1,000, 1,200, 1,400, 1,600, 1,800, 2,000 and 2,400 patients by margins of 3.280, 2.996, 2.766, 2.579, 2.427, 2.299 and 2.050 times the standard error respectively, with the mean difference and standard error corrected for prespecified main effect covariates in a linear model and the critical values following an O’brien-Fleming alpha-spending function. In the Favorable scenario, 79% power is reached at 2,000 patients, with 13%, 27%, 43%, 58%, and 70% power at earlier interims, allowing for earlier detection of a positive effect.

The timing of interim and final analyses will be driven by the number of patients included in the modified intention-to-treat (mITT) analysis subset. This subset consists of patients in whom the intervention can be meaningfully delivered, i.e., those undergoing advanced airway management with continued use of the assigned study valve and manual ventilation.

We conservatively estimate that approximately 25% of randomized patients will not be included in the mITT analysis subset, primarily because ROSC is achieved before advanced airway placement, with a smaller contribution from patients in whom the study valve is removed or ventilation is transitioned to sustained mechanical support before the primary outcome can be meaningfully attributed to the intervention.

### 10.7 Missing data

In the case of missing primary outcomes sensitivity analyses (incorporating best-case and worst-case scenarios and multiple imputation) will be conducted. Secondary outcome parameters will not be imputed.

## 11. ETHICAL CONSIDERATIONS

The institutional review board tasked with the ethical evaluation of the REVIVE-PEEP trial agreed to perform a pilot experiment of simultaneous ethical and Stage-1 peer review at Peer Community In Registered Reports (PCI-RR). This means that all feedback will be considered as one round of review and the author’s reply to both will weigh the two sources of feedback together.

### 11.1 Regulation statement

This study will be conducted according to the principles of the Declaration of Helsinki (75th WMA General Assembly, Helsinki, Finland, October 2024) and in accordance with the Medical Research Involving Human Subjects Act (in Dutch WMO), Medical Treatments Contracts Acts (WGBO) and General Data Protection Regulation (GDPR).

### 11.2 Ethical considerations

The selection of the study population warrants careful consideration, particularly the decision to include all OHCA patients rather than restricting enrolment to those with favorable prognostic characteristics. This choice has important implications for both the ethical acceptability and the scientific validity of resuscitation research. Below, we outline the rationale for our patient selection strategy.

First, prognosis during cardiac arrest is inherently uncertain at the time when interventions must be initiated. Any attempt to prospectively identify patients with a “good prognosis” would rely on imperfect and incomplete information, introducing substantial selection bias.

Excluding patients on the basis of early prognostic indicators risks systematically denying access to potentially beneficial interventions to those who are most vulnerable, thereby violating the ethical principle of justice.

Second, OHCA trials are unavoidably pragmatic in nature, reflecting real-world EMS practice, where CPR quality, response times, and patient characteristics vary widely. An intervention that demonstrates benefit only under idealized conditions may have limited relevance for routine care. Ethically, it is essential that trials evaluate whether an intervention can deliver benefit—or cause harm—across the full spectrum of patients to whom it would be applied if implemented. Selective inclusion of lower-risk patients could overestimate efficacy and obscure harms that might disproportionately affect higher-risk subgroups.

Third, excluding patients with poor prognostic features may mask clinically important heterogeneity in treatment effects. As illustrated in previous trials, interventions may confer benefit in some subgroups while causing harm in others. Identifying such effect modification requires inclusion of the entire OHCA population. Moreover, the intervention under investigation aims to optimize the quality of cardiopulmonary resuscitation —a universally indicated and time-critical emergency procedure during cardiac arrest. While the underlying etiology of cardiac arrest may vary, there is no biological rationale to differentiate between patient groups at the time of inclusion. Ethically, it is unacceptable to adopt or recommend an intervention without understanding whether it worsens outcomes in certain patient groups, particularly those already at highest risk of death.

Finally, from a patient-centered ethical perspective, withholding enrolment from patients deemed unlikely to benefit reinforces a self-fulfilling prophecy, in which poor prognosis becomes a justification for exclusion rather than a target for improvement. Advances in resuscitation care have historically arisen from inclusive research that challenged assumptions about futility.

In summary, inclusion of all OHCA patients in this study is ethically justified because it promotes fairness, preserves scientific validity, enables detection of both benefit and harm across subgroups, and ensures that trial findings are applicable to real-world practice. While the intervention may offer benefit, genuine clinical equipoise remains regarding its effectiveness.

### 11.3 Recruitment and consent

#### 11.3.1 Recruitment procedure

If the dispatch indicates a suspected cardiac arrest, the study resuscitation bag will be brought to the scene. In the standard Advanced Life Support bag used by ambulance services for such cases, the regular resuscitator will have been pre-emptively replaced with a randomly assigned study resuscitator. Upon arrival, the clinical situation is assessed to determine whether CPR is indeed indicated. Randomization is considered to have occurred once the resuscitator bag is used on a patient undergoing CPR, irrespective of airway modality.

#### 11.3.2 Deferred consent

REVIVE-PEEP is conducted in an emergency setting under deferred consent, as defined in Article 6(4) WMO and Article 35 CTR. Subjects suffer an out-of-hospital cardiac arrest and are unconscious at inclusion, so neither prior information nor prior consent can be obtained, and the acute situation leaves no therapeutic window to approach a legal representative beforehand. The study intervention and all study-specific data collection are completed during the prehospital resuscitation; no study-specific procedures or data collection take place in-hospital, and only routinely collected clinical outcome data are added to the dataset. In accordance with the CCMO Memorandum Deferred Consent (7 April 2020), because the investigational procedures — including data collection — are already completed before consent can be sought, the deferred consent obtained afterwards relates to continued participation and to the processing and analysis of the collected data. Strictly speaking, this is therefore not “deferred consent” to the intervention itself, but deferred permission for the continued use of data that has, by necessity, already been collected. For that reason, no fixed deferral window is applied; consent is sought as soon as the clinical situation reasonably allows.

Beyond its legal permissibility, deferring consent is in this setting also ethically preferable to seeking consent at the earliest possible moment. Patients approached during the acute or early intensive care phase may have impaired comprehension and recall, which can compromise the validity of consent.^39–41^ Postponing the consent process until the patient has adequately recovered allows for a more informed and voluntary decision, thereby better safeguarding patient autonomy.

For this reason we prefer, wherever possible, to obtain consent from the patient themselves once they have sufficiently recovered, as the patient is best placed to make an autonomous decision about the use of their own data. Consent is sought from a legal representative only where the patient does not regain sufficient decision-making capacity within a reasonable period. In REVIVE-PEEP this ethical consideration entails no scientific cost: because the intervention and all study-specific data collection are completed during the resuscitation, deferring consent affects only the subsequent processing and analysis of already-collected data, not the conduct of the study. The legal framework and the ethical rationale therefore point in the same direction: towards a consent process that is deferred until the subject, or where applicable their legal representative, is genuinely able to make an informed and voluntary decision.

#### 11.3.3 Consent procedure

Deferred informed consent is primarily sought as soon as feasible during hospital admission:

1. Following the arrest, the subject is admitted to the ICU and undergoes standard post-resuscitation care, including neuroprognostication.
2. After neuroprognostication, the patient information folder (PIF) is provided — preferentially to the subject once they have regained sufficient decision-making capacity. Where the subject has not regained capacity by the time they are discharged or transferred from the ICU — for example to a general ward, another department, or another facility — the PIF is provided to their legal representative instead.
3. The subject or legal representative is given sufficient time to consider the information and to ask questions.
4. Deferred informed consent is subsequently sought, and the signed informed consent form is obtained from the subject or legal representative. Consent and any refusal are recorded in Filemaker. If either the subject or the legal representative declines consent, no study data from that participant will be used for analysis, and any study-specific data already collected will be permanently removed from the study dataset.

All participants are subsequently contacted by telephone as part of scheduled follow-up. This call serves three purposes: it offers the subject the opportunity to opt out of participation if they do not clearly recall having given consent during hospital admission; it offers the opportunity to provide informed consent to subjects who did not give consent during hospital admission — either because they had not yet regained sufficient decision-making capacity, or because they were unwilling to decide immediately after receiving the PIF and preferred more time; and it is used to administer the quality-of-life questionnaire.

In-hospital consent is the preferred route and is pursued in the large majority of cases. However, it is not always possible — for example when subjects are transferred to other ICUs in non-participating hospitals for logistical reasons, or because the five participating ambulance services transport resuscitated patients to more than twenty hospitals across the region, making it infeasible to have a study team member present at every location. To ensure that no subject is left uninformed of their participation and data use as a result of these logistical constraints, the study includes a contingency plan whereby, when deferred consent cannot be obtained during admission:

1. Contact and address details of the subject are obtained from the Personal Records Database (Basisregistratie Personen, BRP), as recorded by the prehospital care provider during prehospital care.
2. The PIF and ICF are sent to the subject’s home address (as registered in the BRP), allowing sufficient time to consider the information.
3. A researcher from ARREST approaches the subject (or, where the subject has not regained sufficient decision-making capacity within a reasonable period, their legal representative) by telephone to answer questions and, if the subject wishes to participate, to obtain deferred informed consent. If the subject is not reached upon the first attempt, two further attempts are made, at most once per week, so that the total period is at least three weeks.

If, after the above attempts, the subject or legal representative cannot be reached, an objection letter is sent to the home address, giving four weeks to object to the use of the already-collected prehospital data, by mail or e-mail. This objection option applies only to the use of data already collected during the resuscitation; it does not constitute consent to participate.

A substantial proportion of subjects will die before consent can be sought. In line with the CCMO memorandum, the data of deceased subjects may be used. The consent procedure described below is illustrated graphically in Figure 7.

**Figure 7.**
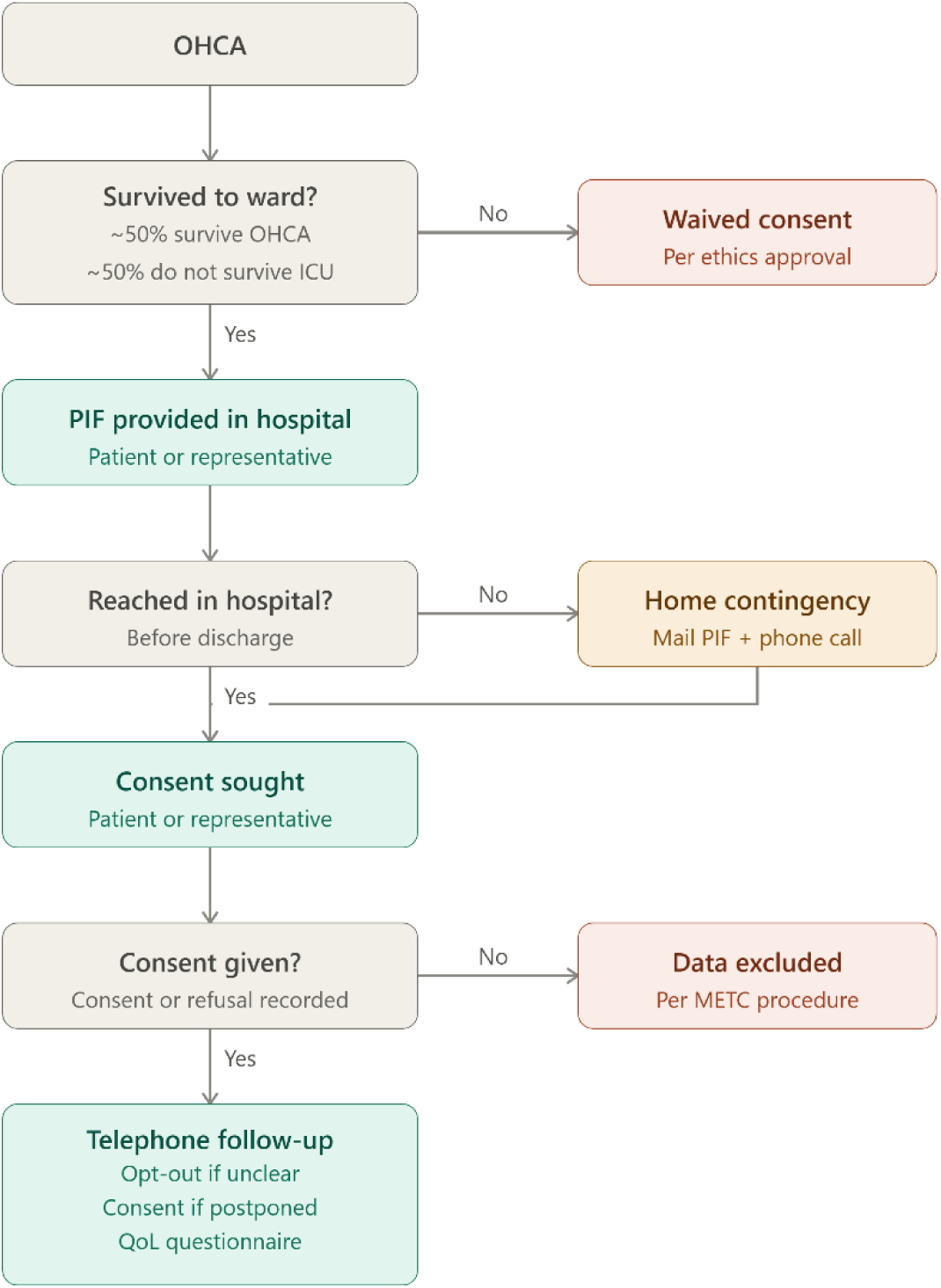
Consent Procedure Flow Chart

### 11.4 Benefits and risks assessment, group relatedness

The intervention is applied during standard emergency care and does not go beyond routine resuscitation procedures. This study can only be conducted in incapacitated patients, as cardiac arrest inherently involves loss of consciousness. It poses minimal risk and no additional burden, as patients are unconscious and cannot experience discomfort. The use of a study resuscitator does not delay or interfere with care. Potential benefits include improved ventilation and possibly better survival or neurological outcomes. There is genuine clinical equipoise regarding optimal ventilation during CPR, and the potential value of this research in improving outcomes justifies the low risk involved.

### 11.5 Compensation for injury

The sponsor/investigator has a liability insurance which is in accordance with article 7 of the WMO. The sponsor (also) has an insurance which is in accordance with the legal requirements in the Netherlands (Article 7 WMO). This insurance provides cover for damage to research subjects through injury or death caused by the study. The insurance applies to the damage that becomes apparent during the study or within 4 years after the end of the study.

## 12. ADMINISTRATIVE ASPECTS, MONITORING AND PUBLICATION

### 12.1 Handling and storage of data and documents

Data for this study will be collected via the AmsteRdam REsuscitation STudies (ARREST) registry, an ongoing, prospective database that captures all resuscitation attempts in the North-Holland province. The ARREST registry has been operational since 2005 and is managed by a dedicated team of data managers and PhD students. Data entry and monitoring follow a detailed data dictionary, aligned with the Utstein OHCA Resuscitation Registry Template, ensuring consistent and high-quality data collection.

Data sources include:

- EMS dispatch centers
- First responders
- AED recordings
- Stryker LIFEPAK 15® & Corpuls® defibrillator data (rhythm strips, thoracic impedance, capnography, 12-lead ECGs, SpO2)
- Ambulance run reports
- Respiratory monitoring devices
- Hospitals, pharmacies, and general practitioners

All collected data are stored in FileMaker Pro 19 and categorized across domains such as system, dispatch, patient, process, and outcomes. Following each cardiac arrest, ambulance personnel routinely submit a standardized case report form and full run sheet to the ARREST team. For this study, the form will be adapted to include the unique identifier of the study-specific resuscitator bag, adherence to protocol, and, if the protocol is not followed, the reason for the deviation.

All subject data will be coded (pseudonymized). The key to the code (linking subject identifiers to study IDs) will be stored separately and securely, with access restricted to authorized ARREST data managers only. Only designated study personnel will have password-protected access to the coded dataset. Identifiable information will never be shared outside the research team.

The standard procedures for data handling and storage within the ARREST registry are documented in the ARREST study protocol (“Onderzoeksprotocol ARREST 2.1”, section 8.1), approved by the METC of Amsterdam UMC (ref. AMC2016-269).

For the REVIVE-PEEP trial specifically, a Data Transfer Agreement was established and signed between the ARREST registry and the regional ambulance services.

All data are stored on secure, access-controlled servers at Amsterdam UMC, with daily encrypted backups. Access to the server and the database is limited to the ARREST team and protected by institutional-level IT security measures in compliance with GDPR. In accordance with European GDPR requirements and Dutch WMO legislation, all subject data will be retained for a minimum of 15 years.

### 12.2 Monitoring and Quality Assurance

Monitoring for this study will be carried out by the Clinical Monitoring Center of the Amsterdam UMC. Research Data Management (RDM), who function as a internal Trusted Third Party, monitors the ARREST data registry by audits and unannounced checks on the data management procedures (as described in the standard ARREST study protocol). A letter from the RDM detailing their involvement, including supervision of the standard operating procedures (“Brief CRU aan BTC_ARREST_20190606”), is available upon request.

### 12.3 Amendments

Amendments are changes made to the research after a favourable opinion by the accredited METC has been given. All amendments will be notified to the METC that gave a favourable opinion.

A ‘substantial amendment’ is defined as an amendment to the terms of the METC application, or to the protocol or any other supporting documentation, that is likely to affect to a significant degree:

- the safety or physical or mental integrity of the subjects of the trial;
- the scientific value of the trial;
- the conduct or management of the trial; or
- the quality or safety of any intervention used in the trial.

All substantial amendments will be notified to the METC and to the competent authority.

Non-substantial amendments will not be notified to the accredited METC and the competent authority, but will be recorded and filed by the sponsor.

### 12.4 Annual progress report

The sponsor/investigator will submit a summary of the progress of the trial to the accredited METC once a year. Information will be provided on the date of inclusion of the first subject, numbers of subjects included and numbers of subjects that have completed the trial, serious adverse events/ serious adverse reactions not described in section 9.2, other problems, and amendments.

### 12.5 Temporary halt and (prematurely) end of study report

The sponsor will notify the accredited METC and the competent authority of the end of the study within a period of 90 days. The end of the study is defined as the finalization of the collection of data from the last patient.

The sponsor will notify the METC immediately of a temporary halt of the study, including the reason of such an action. In case the study is ended prematurely, the sponsor will notify the accredited METC and the competent authority within 15 days, including the reasons for the premature termination.

Within one year after the end of the study, the investigator/sponsor will submit a final study report with the results of the study, including any publications/abstracts of the study, to the accredited METC and the Competent Authority.

### 12.6 Public disclosure and publication plan

This study protocol is a supplement to the Stage-1 Registered Report published at Peer Community In Registered Reports. The study will be registered at a registry of the ICMJE and the study protocol will be submitted for publication before the recruitment of the first patient, in line with the CCMO publication policy and the requirements of the ICMJE. The Stage-1 Registered Report is published by the coordinating investigator under senior and corresponding authorship of the trial statistician dr. Ter Schure. For the protocol publication and result publications, the coordinating investigator will also be the first author, and the principal investigator and dr. Van Schuppen will share last authorship and act as corresponding authors. Additional authorship will follow the guidelines of the International Committee of Medical Journal Editors and will include members of the steering committee and clinical staff as appropriate.

The results of the study will be disclosed unreservedly in a relevant peer-reviewed medical journal. AMBU will be provided with the results after acceptance in a journal for informational purposes, but will not have any influence on the interpretation or presentation of the results.

## 13. STRUCTURED RISK ANALYSIS

### 13.1 Potential issues of concern

#### a. Level of knowledge about mechanism of action

PEEP is a well-established intervention in all ventilated patients in elective anesthesia and critical care, primarily used to maintain alveolar recruitment and prevent atelectasis. Its inconsistent use during CPR is an exception, given its widespread application in ventilated patients across various patient populations.

There is a biologically plausible mechanism supporting the use of PEEP during CPR. Both animal and clinical studies consistently demonstrate severe atelectasis during CPR, which PEEP helps to alleviate by improving oxygenation. While arterial blood pressure is typically maintained in most animal studies, one study suggests that PEEP levels greater than 10 cm H_2_O slightly reduce oxygen delivery. This reduction is derived from the calculation of cardiac output multiplied by the oxygen-carrying capacity of the blood, indicating potential hemodynamic effects at higher PEEP levels.

#### b. Previous exposure of human beings with the medical device with a similar biological mechanism

The primary mechanism of action of manual and mechanical PEEP is similar, with both serving to maintain alveolar recruitment and improve oxygenation. However, there is a slight nuance in how pressure is generated. Manual PEEP, delivered via a passive valve, relies on the exhalation to generate pressure, while mechanical PEEP is actively generated by a ventilator, which adjusts for dynamic changes in airway pressure during CPR.

During CPR, chest compressions produce a dynamic expiratory pressure. During decompression, the pressure drops due to a relative underpressure in the airway when using a passive valve for manual PEEP. A mechanical ventilator, however, can actively adjust to this pressure difference to maintain the desired PEEP level.

This dynamic difference in pressure generation might favor manual PEEP during decompressions, as the pressure drop can facilitate venous return by promoting blood flow back to the heart. On the other hand, higher intrathoracic pressures during decompression, such as those created by mechanical ventilators, might reduce venous return and possibly impair circulation.

Despite the slight mechanistic difference in how PEEP is generated, the majority of its effects are expected to be similar between the two methods. Both aim to increase residual lung volume by limiting passive expiration and providing active limitation as a result of chest compressions. Both manual and mechanical PEEP are used in clinical practice, and while the European Resuscitation Council guidelines recommend setting PEEP between 0 and 5 cm H_2_O during CPR, in actual clinical settings, a broader range of PEEP is commonly applied by physicians.

#### c. Can the primary or secondary mechanism be induced in animals?

Animal studies demonstrate that oxygenation can be improved with the application of PEEP compared to ZEEP during CPR. These studies show enhanced oxygen delivery and better alveolar recruitment, leading to improved gas exchange during resuscitation. While the effects on hemodynamics may vary depending on the study design and outcome measures, the primary mechanism—improved oxygenation via increased residual lung volume—has been reliably induced in animal models.

#### d. Selectivity of the mechanism to target tissue in animals and/or human beings

Not applicable, PEEP is a mechanical intervention that primarily targets the lungs.

#### e. Analysis of potential effect

Based on preclinical and clinical studies, the highest tolerable PEEP dose is 10 cm H_2_O, with no significant hemodynamic effects. Clinical studies suggest the optimal PEEP dose during CPR is ≥8 cm H_2_O, with a tolerance range of ±2 cm H_2_O, balancing effectiveness and safety within the maximum tolerated range.

#### f. Pharmacokinetic considerations

Not applicable.

#### g. Study population

The study population consists of patients experiencing OHCA not related to trauma or drowning, which represents the ultimate consequence of a life-threatening disease. These patients are critically ill, with unstable conditions, and require emergency health care.

#### h. Interaction with other products

Not applicable.

#### i. Predictability of effect

Extrapolating animal data to clinical scenarios, particularly in cardiac arrest, is inherently challenging. While a significant portion of the evidence comes from animal studies, the purported benefits observed in healthy, anesthetized animals in a controlled lab setting often fail to translate into clinical practice where CPR occurs in chaotic, prehospital settings where ventilation is often inadequate. Animal models are demonstrating biological plausibility, but their findings must be interpreted cautiously, with a strong emphasis on clinical corroboration through well-designed studies such as REVIVE-PEEP.

While observational studies often suggest large effect sizes, most interventions during cardiac arrest have failed to show significant benefit in randomized controlled trials.

Therefore, we maintain a modest expectation for the effect of PEEP, recognizing that the larger effect sizes seen in observational studies have generally not been borne out in randomized trials, and we have taken this into account when determining the MCID.

#### j. Can effects be managed?

The primary effect that may require management is the occurrence of pneumothorax, a known risk with CPR but that might be exacerbated. However, it is part of the standard diagnostic approach to rule out pneumothorax in every cardiac arrest case, and treatments are readily available if it occurs.

### 13.2 Synthesis

The risks associated with participation in this study are minimal and are outweighed by the potential benefits, making them acceptable within the context of this trial. While there is a theoretical risk related to defibrillation efficiency and the occurrence of pneumothorax, there is no clinical data to suggest that these risks are likely to occur or have significant impact.

From a clinical standpoint, the potential benefits—such as improved oxygenation, enhanced ventilation, and an increased likelihood of successful resuscitation and favorable neurological outcomes—significantly outweigh the theoretical risks. Therefore, there is genuine equipoise between the treatment groups in terms of both risks and benefits.

To ensure participant safety, the DSMB will oversee interim analyses, providing an additional layer of oversight to identify and address any emerging risks. This continuous monitoring will allow for the early detection and management of any adverse effects, further ensuring the acceptability of the study’s risks. The uncertainty surrounding the optimal use of PEEP and its impact on clinical outcomes is central to this study, and this equipoise justifies the continuation of the research.

## Data Availability

All data produced will be made available in the Replication package (for reproducibility and replication) at ResearchEquals: https://doi.org/10.53962/p8t5-wzwr

## LIST OF ABBREVIATIONS AND RELEVANT DEFINITIONS

AE: Adverse Event
AED: Automatic external defibrillator
ALS: Advanced Life Support
ARREST: AmsteRdam REsuscitation Studies
CONSORT: Consolidated Standards of Reporting Trials
BVM: Bag-valve-mask
CCMO: Central Committee on Research Involving Human Subjects; in Dutch: Centrale Commissie Mensgebonden Onderzoek
CPR: Cardiopulmonary resuscitation
CRF: Case report form
DSMB: Data Safety Monitoring Board
ETT: Endotracheal tube
GDPR: General Data Protection Regulation; in Dutch: Algemene Verordening Gegevensbescherming (AVG)
IB: Investigator’s Brochure
ILCOR: International Liaison Committee on Resuscitation
MCID: Minimally Clinically Important Difference
METC: Medical research ethics committee (MREC); in Dutch: medisch-ethische toetsingscommissie (METC)
MRS: Modified Rankin Scale
OHCA: Out-of-hospital cardiac arrest
PEEP: Positive end-expiratory pressure
ROSC: Return of spontaneous circulation
(S)AE: (Serious) Adverse Event
SAD: Supraglottic airway device
SPC: Summary of Product Characteristics; in Dutch: officiële productinformatie IB1-tekst
Sponsor: The sponsor is the party that commissions the organisation or performance of the research. A party that provides funding for a study but does not commission it is not regarded as the sponsor, but referred to as a subsidising party.
SUSAR: Suspected Unexpected Serious Adverse Reaction UW-mRS Utility-weighted modified Rankin Scale
WMO: Medical Research Involving Human Subjects Act; in Dutch: Wet Medisch-wetenschappelijk Onderzoek met Mensen
ZEEP: Zero end-expiratory pressure

## 15 APPENDIX 1

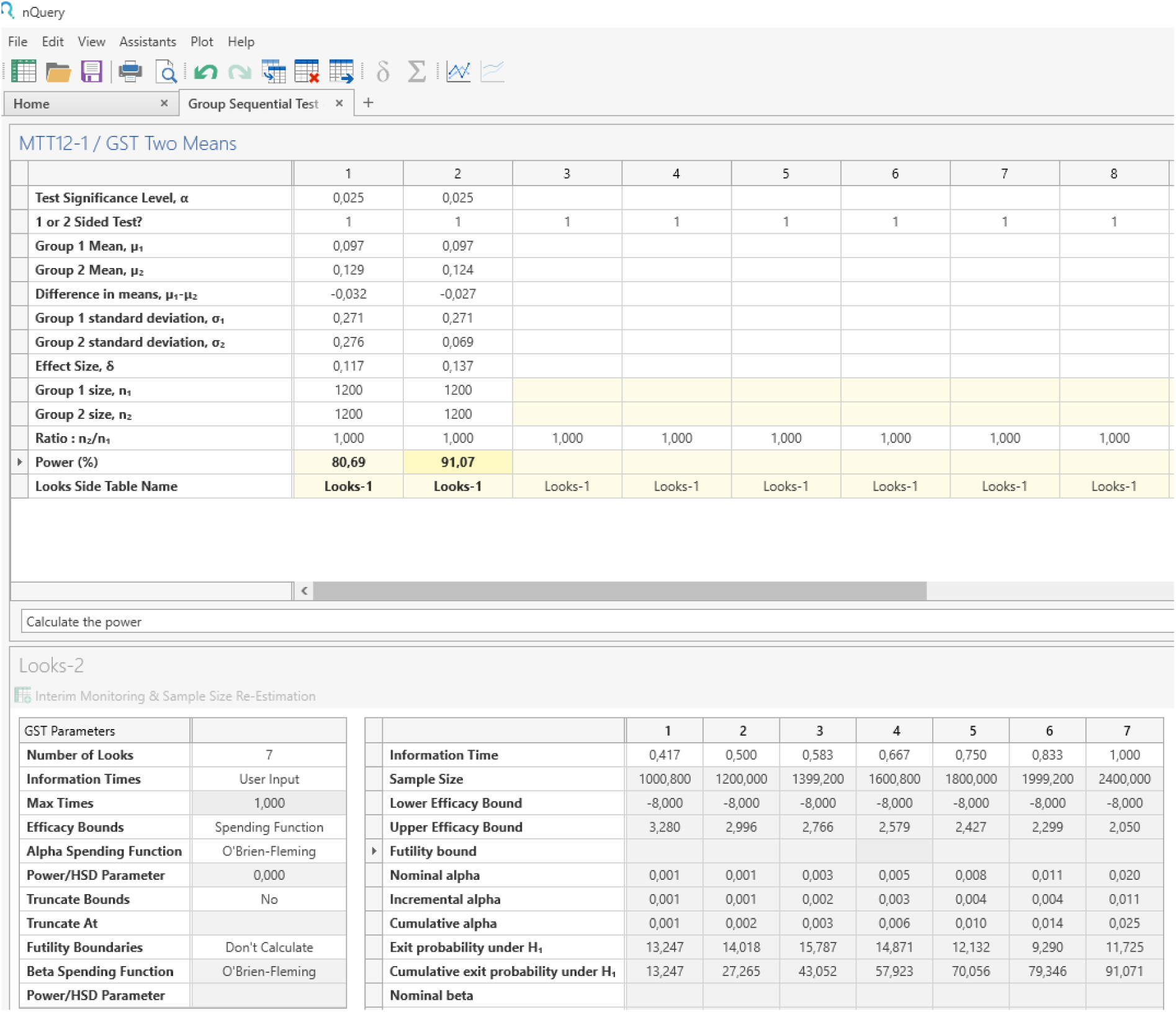

